# Feature analysis of joint motion while reaching the occiput in patients with mild hemiplegia: a cross-sectional study

**DOI:** 10.1101/2023.11.15.23298611

**Authors:** Daigo Sakamoto, Toyohiro Hamaguchi, Naohiko Kanemura, Takashi Yasojima, Keisuke Kubota, Ryota Suwabe, Yasuhide Nakayama, Masahiro Abo

## Abstract

This cross-sectional study aimed to clarify the kinematic characteristics of reaching the occiput in patients with mild hemiplegia. Ten patients with post-stroke hemiplegia who attended the Department of Rehabilitation Medicine of the Jikei University Hospital and met the eligibility criteria were included. Reaching motion to the back of the head by the participants’ paralyzed and non-paralyzed upper limbs was measured using three-dimensional motion analysis, and the motor time, joint angles, and angular velocity were calculated. Multivariate analysis of covariance was performed on these data. After confirming the fit to the binomial logistic regression model, the cutoff values were calculated using the receiver operating characteristic curve. The cutoff values for the movement until the hand reached the back of the head were 1.6 s for the motor time, 55° for the maximum shoulder joint flexion angle, and 145° for the maximum elbow joint flexion angle. The cutoff values for the movement from the back of the head to the hand being returned to its original position were 1.6 s for the motor time, 145° for the maximum elbow joint flexion angle, 53°/s for the maximum angular velocity of shoulder joint abduction, and 62°/s for the maximum angular velocity of elbow joint flexion. The numbers of clusters were three, four, and four for the outward non-paralyzed side, outward and return paralyzed side, and return non-paralyzed side, respectively. The findings obtained by this study can be used for practice planning in patients with mild hemiplegia who aim to improve the reaching motion to the occiput.

## Introduction

Stroke affects 14 million individuals annually worldwide, and 80 million people live with the aftereffects of stroke [1]. Motor paralysis, a sequela of stroke, occurs in approximately 80% of patients [2]. As motor paralysis of the upper limbs and fingers limits patients’ activities of daily living (ADLs) and decreases their quality of life (QOL), patients are provided with continuous rehabilitation to improve motor paralysis [3,4].

Reaching the occiput is an important exercise for achieving rehabilitation. Reaching is a fundamental movement of the upper limb, with the goal of reaching the target point [5]. Although there are various target points for reaching, such as a space or an object on a desk, a reach whose target point is one’s body has a direct influence on self-care performance. The movement of the hand to the back of the head is included in movements for grooming, such as washing, tying hair, and putting on and taking off clothes, ornaments and hats [6]. Improving the patient’s appearance positively affects their QOL through social participation, such as going out and socializing. Reaching the back of the head requires a wide range of motion and coordinated joint movement of the shoulder and elbow joints and is a difficult task for patients with motor paralysis; therefore, even if the patient’s motor paralysis is mild, smooth movement requires practice [7,8]. Therefore, therapists monitor changes in patients owing to exercise and treatment and provides further treatment based on the evaluation.

Clinically, the arm function test, manual function test, and action research arm test (ARAT) are used for upper extremity function assessment to observe the reach to the occiput include [9–11]. In these tests, the examiner observes whether the patient can perform the task and the reaching position of the patient’s hand at the end of the motor limb; the test is scored using an ordinal scale. However, as the human body has redundant degrees of freedom, multiple combinations of joint motions exist, even when the final hand position is the same [12].

Specifically, in patients with hemiplegia after stroke, synergistic patterns may emerge, and compensatory movements may be used [13]. In patients with motor paralysis, even if reaching the occiput is possible, the motion trajectory may be prolonged and the motion time may be delayed. The scores obtained using the clinically used evaluation methods alone are difficult to refer to when planning exercises to change a patient’s joint movement patterns or shorten the motor time of exercises.

Three-dimensional motion analysis is used for analyzing upper limb movement characteristics in patients with stroke [14]. In this method, markers are attached to landmarks on the body and upper limb movements are recorded using an infrared camera and analyzed. Studies have been conducted to analyze the joint motion and motion time of patients with stroke using three-dimensional motion analysis [15–18]. In these studies, reaching a target placed in front or on a desk is occasionally set as the measurement task. However, it is difficult to utilize the obtained results for practice because of the discrepancy between the results and the upper limb movements in daily activities [19]. As the tasks are measured in a manner that is close to daily life situations, the results can be easily applied to the treatment of patients [20,21]. Several studies have evaluated healthy participants and patients with orthopedic diseases in terms of their ability to reach the occipital region; however, the kinematic characteristics of patients with stroke with mild motor paralysis have not been verified [7,8,22]. The data on the kinematic characteristics of patients with stroke with regard to reaching backward to the occiput would provide a useful basis for devising a practice method for those who aim to acquire ADLs, including reaching backward to the occiput. When upper limb motor tasks are measured using a three-dimensional motion analyzer, the motions of the paralyzed and non-paralyzed sides should be compared [23]. The movements of the non-paralytic upper limb in patients with hemiplegia after stroke differ from those of normal participants; however, the non-paralytic upper limb, which does not show obvious functional impairment, can be used in clinical situations for comparison with the paralytic upper limb [23,24]. In a recent study, accelerometers, gyroscopic sensors, and magnetic sensors were attached to the bodies of patients with stroke. The results of four types of forward reaching were compared between the paralyzed and non-paralyzed upper limbs using three-dimensional motion analysis to obtain evaluation values for patients with mild hemiplegia based on the motor time and joint angle of the non-paralyzed side of the upper limb [25]. Three-dimensional motion analysis is clinically useful because the motion of the non-paralyzed upper limb is referenced, and the joint motion patterns of the paralyzed upper limb can be used to devise isolation exercises to be practiced [19].

Based on the above discussion, in this study, we aimed to clarify the kinematic characteristics of patients with mild hemiplegia in the chronic phase while reaching the occipital region using three-dimensional motion analysis. Our results will provide a basis for devising motor targets and effective exercises to reach the occiput in patients with posterior upper limb hemiplegia.

## Materials and methods

### Study design

In this cross-sectional study, kinematic data of patients with post-stroke hemiplegia were obtained while reaching the hand to the back of the head, and the patient’s paralyzed and non-paralyzed upper limbs were compared.

### Ethical considerations

All patients provided written informed consent to participate in this study. This study was approved by the Ethics Committee of the Jikei University School of Medicine (approval number: 22-061-6238).

### Participants

Patients with post-stroke hemiplegia who attended the Department of Rehabilitation Medicine of the Jikei University Hospital between May 1 to October 30, 2020, cases where at least 6 months had passed since the onset of stroke, patients aged ≥20 years, and those with a total score of ≥47 points in the Fugl–Meyer assessment of the upper extremity (FMA-UE) or with a gross movement score of ≥6 points in the ARAT, were included. The 47 points achieved in the FMA-UE is the cutoff for mild motor paralysis in the severity classification of motor paralysis reported by Woodbury et al. [26]. The gross movement in the ARAT includes a task to reach the back of the head, and 6 points of gross movement is the threshold for patients who can perform this movement [10]. The exclusion criteria were as follows: cases with a paralyzed hand not reaching the external occipital ridge with automatic movements; presence of a central nervous system disease other than stroke, orthopedic disease, mental disorder, higher brain dysfunction, dementia, visual field disorder, and ataxia at diagnosis; subluxation of the shoulder joint; pain in the joints of the upper limb or fingers during exercise; presence of a significant limitation in the joint range of motion in the upper limb; and completion of the occupational therapy intervention. Patients who met the study eligibility criteria but did not meet the exclusion criteria were asked to participate in the study, and those who provided consent were considered participants.

### Sample size

The minimum sample size was calculated to be eight patients using G*Power 3.1 (University of Dusseldorf, Dusseldorf, Germany). To calculate the cutoff values of the kinematic data by performing binomial logistic regression analysis using the paralyzed and non-paralyzed upper limbs as nominal variables in patients with hemiplegia, data from previous studies [27], in which the primary assessment was the joint angle, were referenced. The sample size was calculated by setting the difference from the constant (binomial test, one sample case). For calculating the required sample size, the effect size was 0.4, α was 0.05, power (1-β) was 0.8, and constant proportion was 0.5.

### Survey period

The acquisition of patients’ medical information, clinical evaluation, and measurement of motor tasks began on October 1, 2020, and ended on October 1, 2021.

### Experimental procedure

The examiner affixed infrared reflective markers at 35 points on the participant’s body according to the Plug-in Gait marker model (Fig 1). Infrared reflective markers were applied by a single examiner similarly in all participants according to the following procedure: the examiner made the participants sit on a backless chair with their elbow joints fully extended, their forearms in the middle, and their upper limbs drooped. The examiner adjusted the height and position of the chair such that the participants’ forearms and fingers did not touch the chair, and the flexion angles of the knee and hip joints were 90°. The participants sat on a height-adjusted chair with their feet shoulder-width apart and both soles of their feet on the floor. Six thermal imaging cameras were placed on the ceiling of the room, and a landmark was placed 5 m away from the participants (Fig 2).

**Fig 1.**
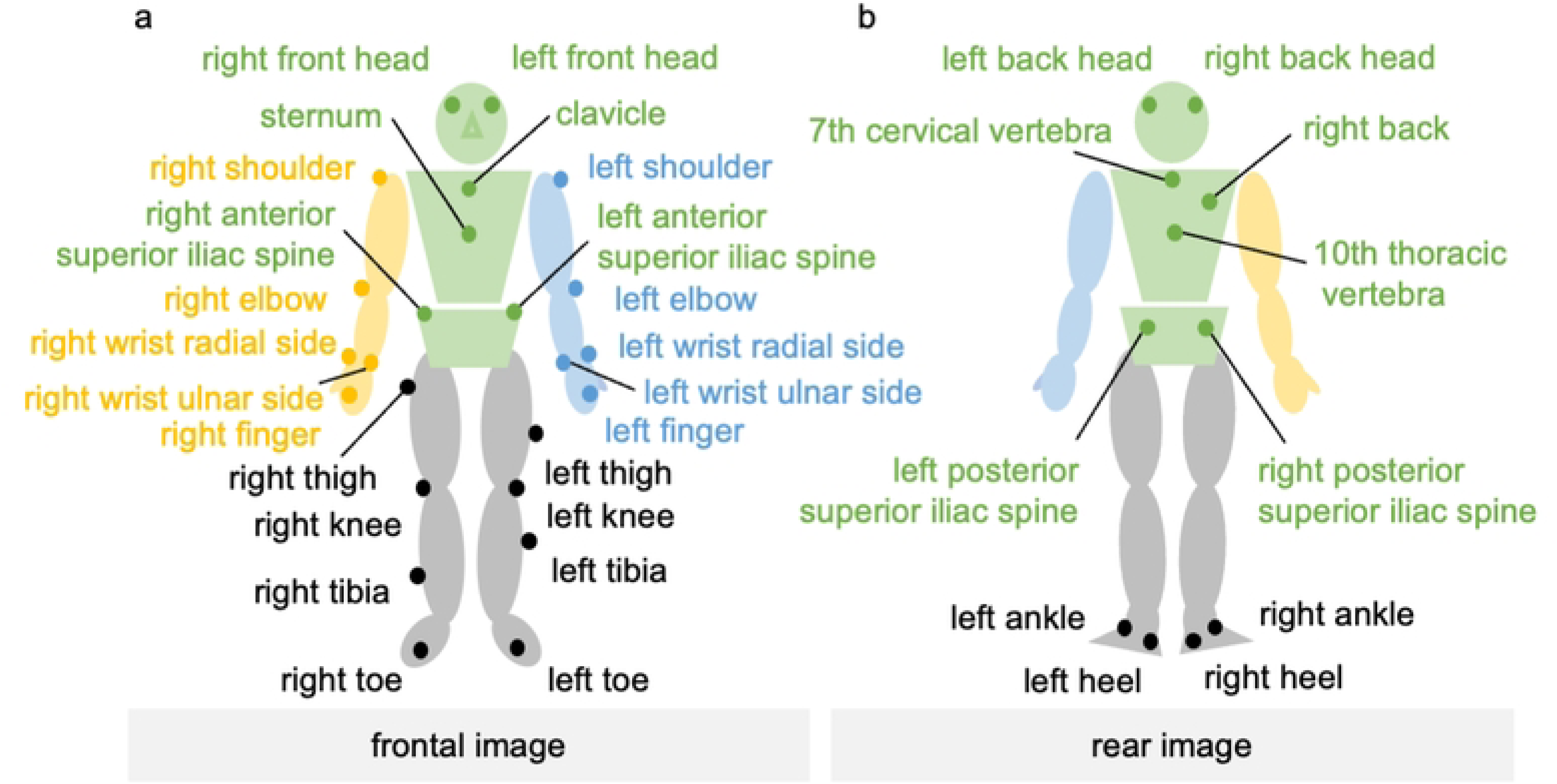
Attachment positions of the markers. (a) Frontal image and (b) rear image.

**Fig 2.**
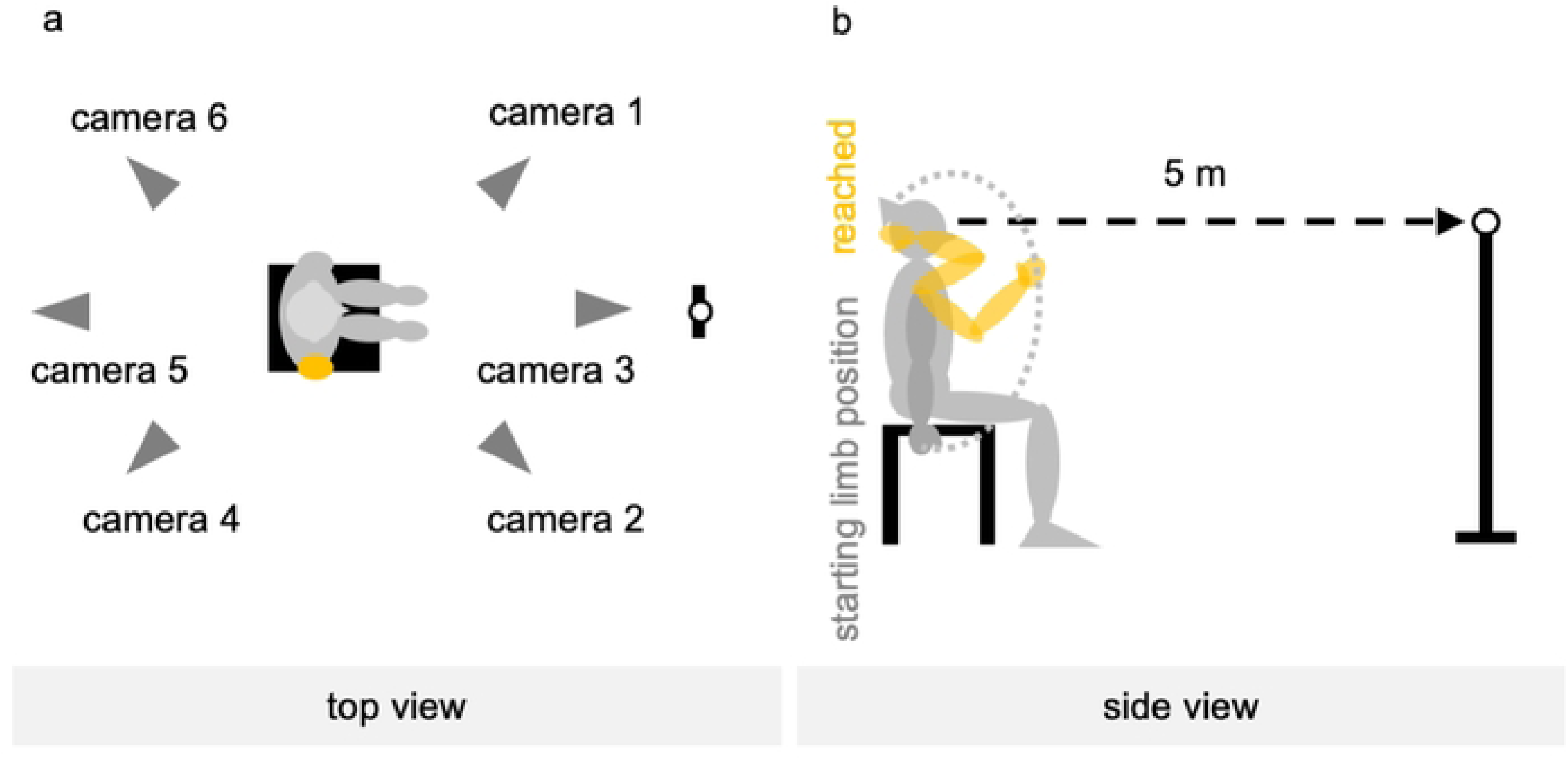
Measurement environment and starting limb position. (a) Top view and (b) side view.

Markers were affixed at 35 points on the bodies of the participants. On the head, they were placed at four bilateral anterolateral and posterolateral points; on the trunk, they were placed at five points on the upper sternal body, lower sternal spine, seventh cervical vertebra, 10th thoracic vertebra, and right posterior back; and on the pelvis, at four points on the bilateral anterior and posterior superior iliac spines. On the upper limbs, they were affixed at 10 points on the bilateral upper acromion, lateral olecranon, radial and ulnar eminence, and the head of the second metacarpal. On the lower limbs, they were affixed at 12 points on the bilateral femur, lateral patella, tibia, calcaneus, external calcaneus, and head of the second metatarsal bone.

### Reaching task

The starting position of the upper limb on the measurement side was set with the elbow in full extension, forearm in mid-extension, and fingers in extension, with the forearm and fingers not in contact with the chair. The upper limb on the non-tested side was also placed in the same position. If the starting position was difficult to achieve owing to paralysis and muscle tone, the participants were reminded to relax the muscles of the upper limbs and fingers, and the upper limbs were allowed to droop as much as possible (Fig 2b).

The reaching task involved placing the palmar aspect of the hand (the palmar aspect of the second metacarpal) in contact with the center of the external occipital ridge. The examiner provided the following verbal instructions to all participants: “touch the center of the occiput with the palm of the hand”; “return the upper limbs to the starting posture; maintain the head, neck, and trunk as still as possible while looking at the landmarks placed in front during the measurement”; and “move the arms as usual without any particular awareness of speed.” Flexion of the fingers during the reaching motion was allowed. The examiner asked the participants to perform the exercise several times and checked whether they understood the instructions correctly.

Measurements were performed on the paralyzed and non-paralyzed sides, in that order, five times each for a total of 10 measurements (Fig 3). A 1-min rest period was allowed between the measurements. Before each measurement, the examiner verified whether the participants were in the correct starting position. The order of the measurements did not change, and all participants underwent measurements in the same manner. During the rest period, the participants were allowed to stretch the muscles by themselves but were not allowed to receive any therapeutic intervention from the therapist.

**Fig 3.**
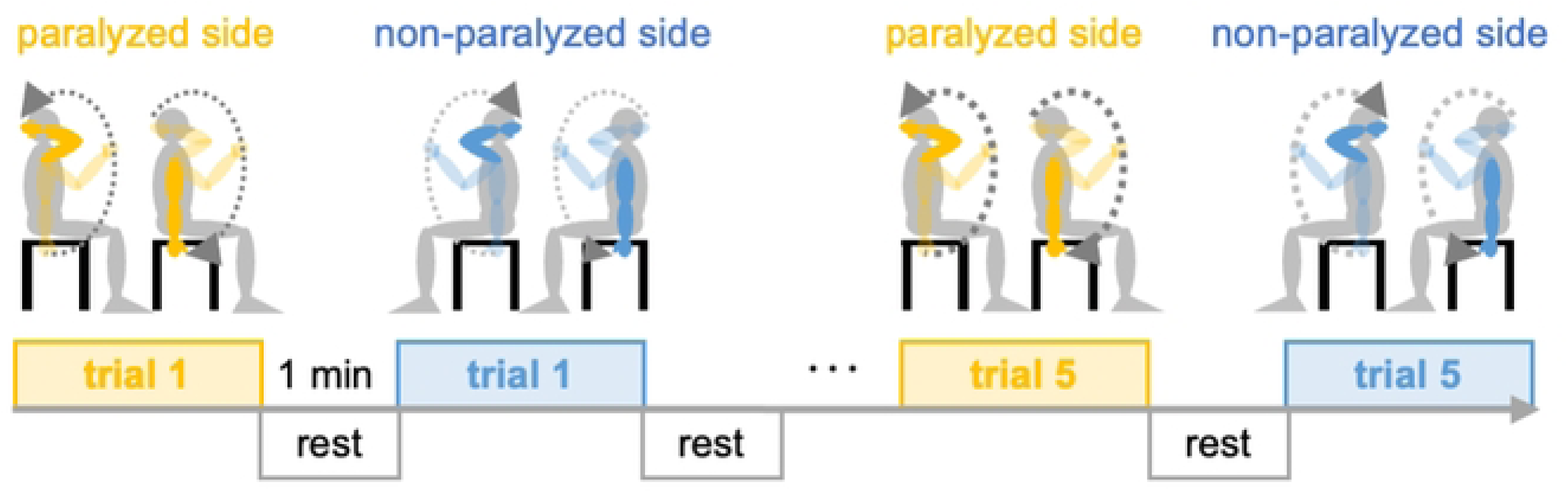
Measurement sequence.

### Data acquisition and analysis

The reaching task was analyzed using an optical three-dimensional motion analyzer (Vicon motion system; Oxford, UK). Data were recorded at a sampling rate of 100 Hz using six infrared cameras mounted on the ceiling of the examination room. The displacement information for each marker was compiled in three dimensions using parallax images from the six cameras, converted into positions (x, y, and z) in the same virtual space, and recorded on a personal computer for analysis. The motion was divided into an outward motion from the starting position until the hand reached the occiput (outward motion) and a return motion from the point when the hand reached the occiput until it returned to its original position (return motion). The onset of the outward and return motions was defined as the time when the position data of the index finger marker in the three-dimensional space changed continuously for 0.2 s, and the end of the movement was defined as the time when the position data of the index finger marker recorded the same value continuously for 0.2 s [28].

A whole-body rigid-body linkage model was constructed from the acquired reflective marker information using the Plug-in gait model specified by the Vicon motion system [29]. The flexion and abduction angles of the shoulder joint as well as the flexion angle of the elbow joint were calculated using the Eulerian method, in which the joint angles are calculated from the distal body segment coordinates in motion relative to the proximal body segment coordinates. The angular velocity was calculated by dividing the change in the joint angle by the time of motion. The displacements of the markers attached to the sternal pattern and occiput on the same side as the measured upper limb during the measurement were calculated using the position data in three-dimensional space. The mean, standard deviation, and maximum and minimum values of the joint angles and angular velocities of shoulder flexion and abduction and elbow flexion, and displacements of the sternal pattern and occipital markers were calculated separately for outward and return motions.

### Clinical evaluation

The FMA was used to assess the motor paralysis of the participants. The FMA is a comprehensive evaluation battery that tests motor function, balance, sensory function, joint range of motion, and the degree of joint pain in patients with post-stroke hemiplegia [30]. Upper and lower extremity motor function (FMA of the lower extremity [FMA-LE]) items can be used in excerpts and assess isolated movements in accordance with the recovery phase of motor paralysis. The FMA-UE is scored on a 66-point scale, and the FMA-LE is scored on a 34-point scale on a 3-point ordinal scale. The FMA-UE has been reported to classify the severity of motor paralysis, and the severity of motor paralysis was investigated in the present study based on the classification given by Woodbury et al. [26]. The ARAT and Box and Block Test (BBT) were used to assess the participant’s ability to manipulate objects. The ARAT is an upper limb function assessment tool, which was developed based on the upper extremity test [31]. The ARAT consists of grasp, grip, pinch, and gross movement subtests and is scored on a 4-point ordinal scale on a 57-point scale [10]. The BBT is used to evaluate hand dexterity. The task is to move 100 blocks from one compartment of a box to the opposite compartment one by one across a partition [32,33]. In this test, the number of blocks moved per minute is measured. The modified Ashworth scale (mAs) was used to assess the muscle tone of the study participants.

The mAs is used to evaluate spasticity, a symptom of abnormal muscle tone in central nervous system diseases [34]. In this test, the resistance to rapid movement of the joints in an alternating manner is evaluated on a 6-point scale. In this study, the biceps brachii muscle of the paralyzed side of the participants was evaluated. The Berg balance scale (BBS) and the functional reach test (FRT) were used to assess the participant’s balance function. The BBS is used to evaluate the functional balance ability and is useful as an indicator of walking independence and fall prediction [35,36]. The test is scored on a 5-point ordinal scale on a 56-point scale. The FRT is a balance ability assessment that measures the distance of the forward reach of the upper limb in the standing position without changing the basal plane of support [37]. In this work, the FRT was performed three times, and the mean value was calculated. The Semmes–Weinstein monofilament test (SWT) and the thumb search test (TST) were used to evaluate the sensory function of the participants. The SWT is used to examine static tactile sensations related to object properties, discrimination ability, and sustained grasping [38]. In this test, five types of nylon filaments with different diameters are used to stimulate the skin, and the participant’s responses are evaluated. The TST is used to evaluate the joint localization of the upper limb [39]. This test integrates the proprioceptive information of each joint of the upper limb and evaluates the perception of thumb position in space on a 4-point ordinal scale. The Barthel Index (BI) was used to assess participants’ daily functioning in this study. The BI is used to assess the level of independence in performing ADLs [40]. The test consists of 10 items, and the degree of assistance is evaluated using a 4-point ordinal scale. If the patient independently performs all items, the score is 100 points; if the patient requires full assistance for all items, the score is 0 points.

### Participant characteristics

Basic and medical information regarding the participants, including sex, age, height, body mass index (BMI), stroke type, post-stroke duration, and paralytic side, was collected from their medical records. The Edinburgh hand test was used to examine the handedness of the participants [41]. This test consists of 10 questions, in which the participants are asked which hand they use to perform ADLs. A positive index indicates right-handedness, whereas a negative index indicates left-handedness. In this study, the dominant hand before the stroke and the current dominant hand were investigated.

### Statistical analysis

Multivariate analysis of covariance was conducted to test the hypothesis that the kinematic characteristics of reaching the back of the head in patients with mild hemiplegia in the chronic phase differ between the paralyzed and non-paralyzed upper limbs. The dependent variable was the measured side of the reaching task, and the independent variables were motor time, maximum values of joint angles, and angular velocities of shoulder flexion and abduction and elbow flexion. The covariates included sex, age, BMI, time since stroke onset, maximum displacement of the sternal pattern, and occipital markers. Statistical analyses were conducted separately for the outbound and return motions. Binomial logistic regression analysis was conducted on the features fitted to the model for the paralyzed and non-paralyzed sides using multivariate analysis of covariance. After confirming the fit of the binomial logistic regression model, Youden’s index was calculated using the receiver operating characteristic (ROC) curve, and the cutoff value to discriminate between the paralyzed and non-paralyzed sides was calculated. The calculated cutoffs, especially the area under the curve (AUC) values, were compared using the Delong test [42]. The dependent variables were the paralyzed and non-paralyzed sides, and the independent variables were the characteristics that showed significant differences between the paralyzed and non-paralyzed sides in the multivariate analysis. The covariates included sex, age, BMI, post-onset period, and the maximum displacement values of sternal pattern markers and occipital markers. For these analyses, jamovi version 2.2.1 (https://www.jamovi.org) was used.

Pattern identification using random forest clustering was performed to analyze the pattern of changes in the joint angle on the paralyzed and non-paralyzed sides of reaching the occipital region. Random forest clustering is an algorithm that divides the data into several clusters, virtually partitioning the data such that each observation belongs to only one group. The clustering method is an unsupervised method that uses the random forest algorithm, wherein the dependent variable *y* is set as *T*, the number of decision trees, and the machine is trained with each motor data input as unlabeled data with and without motor paralysis (Equation 1). The random forest algorithm generates a proximity matrix that estimates the distance between the observations based on the frequency of observations ending at the same leaf node (Equation 2). These data consist of continuous variables [43–45].

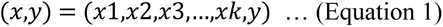

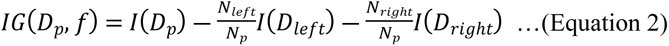

In Equation 1, *x* is the reference dataset and *y* is the dependent variable. In Equation 2, *Dp* is the parent dataset, *f* is the number of explanatory variables, *N* is the depth of the decision tree, *D_left_* is the left child node from the parent node, *D_right_* is the right child node from the parent node, and *I* is the impurity. The variables used were the motor time and shoulder flexion, shoulder abduction, and elbow flexion angles. Each variable *C* was assigned a discrete-valued class label, that is, C0, C1, C2,......, Cn, using random forest clustering. The motor patterns were clustered for four conditions: outward and return motions on the paralyzed and non-paralyzed sides of the reach to the occipital region. The number of clusters was determined using the elbow method. The elbow method plots the sum of squares of the intra-cluster error for each cluster and considers the point where the value sharply decreases to be the optimal number of clusters [46]. The clustering structure of these four conditions is illustrated and is considered a unique pattern underlying the motion data. JASP version 0.16 (https://jasp-stats.org/) was used to identify these patterns. The statistical significance level was set at 5%.

## Results

### Participants

In total, 88 patients with chronic stroke had a history of intervention in the previous 6 months. Among these patients, three were aged <20 years, and 60 had FMA-UE or ARAT scores below the inclusion criteria. One patient was diagnosed with higher brain dysfunction, two had ataxia, two had shoulder subluxation, and seven completed the occupational therapy intervention. Of the 88 patients, 75 were excluded and 13 met the inclusion criteria set for the study. These 13 patients were asked to participate in the study; three patients who did not agree to participate were excluded; therefore, 10 patients were included in the final analysis (Fig 4).

**Fig 4.**
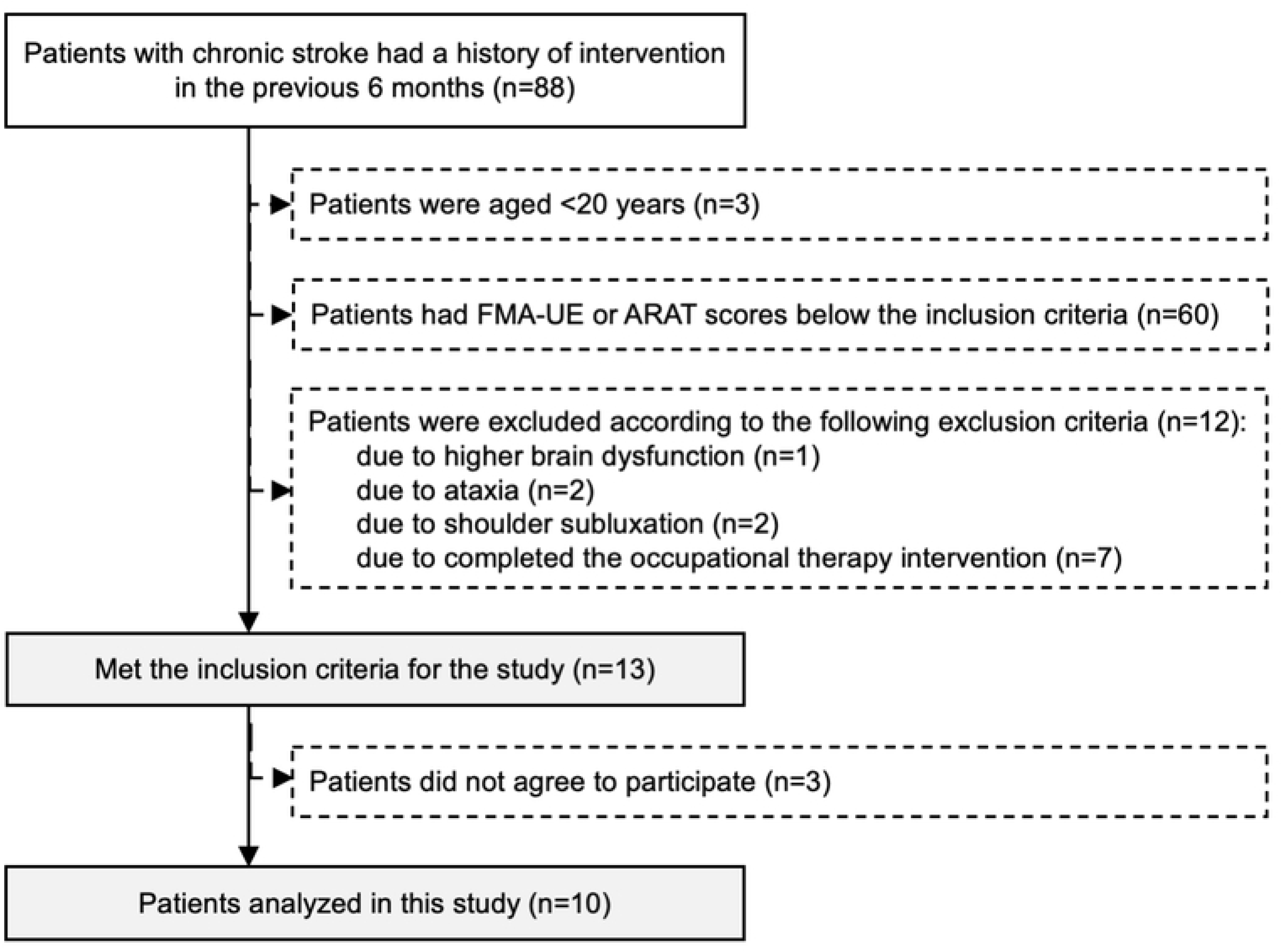
Patient selection procedure.

### Descriptive data

The characteristics of the participants and the survey results are presented in Table 1. In total, 10 participants were included, of whom three were female and seven were male individuals. All the participants had right hemiplegia. The right hand was dominant in 10 participants before the onset of FMA-UE and changed to the left hand in five of 10 participants after the onset of FMA-UE. The severity of FMA-UE was moderate in one patient and mild in nine patients. One participant classified as moderate met the eligibility criterion of at least 6 points for gross movement on the ARAT. The median scores of all the participants were 34 points on the ARAT and 24 points on the paralytic side in the BBT. Eight and two patients had mAs scores of 1 and 1+, respectively. None of the patients showed severe spasticity. The FMA-LE score, BBS score, and FRT were 28 points, 55 points, and 33 cm, respectively. The participants had good lower limb, trunk, and balance functions. The SWT was normal in seven participants and lowered in three, while the TST was normal in nine participants and lowered in one; there was no significant decrease in the superficial or deep sensation in the participants. All participants had a BI score of 100 points and independently performed all daily activities.

**Table 1.** Clinical characteristics of the patients.

| Characteristics |  | Female | Male | All |
| --- | --- | --- | --- | --- |
| Participants |  | 3 (30) | 7 (70) | 10 (100) |
| Age (years) |  | 50 [35–51] | 46 [46–55] | 48 [45–51] |
| Height (cm) |  | 154 [152–159] | 170 [169–173] | 169 [164–171] |
| Weight (kg) |  | 51 [51–56] | 72 [66–77] | 66 [55–73] |
| BMI (kg/m<sup>2</sup>) |  | 23 [21–24] | 25 [22–26] | 25 [20–25] |
| Time from onset (months) |  | 59 [37–83] | 53 [38–78] | 54 [34–91] |
| Diagnosis | CI | 3 (100) | 3 (43) | 6 (60) |
|  | ICH | 0 (0) | 4 (57) | 4 (40) |
| Paralyzed side | Left | 0 (0) | 0 (0) | 0 (0) |
|  | Right | 3 (100) | 7 (100) | 10 (100) |
| Dominant hand |  |  |  |  |
| Before onset | Left | 0 (0) | 0 (0) | 0 (0) |
|  | Right | 3 (100) | 7 (100) | 10 (100) |
| After onset | Left | 2 (67) | 3 (43) | 5 (50) |
|  | Right | 1 (33) | 4 (57) | 5 (50) |
| FMA-UE |  | 56 [52–57] | 57 [55–60] | 57 [55–58] |
| FMA-UE severity |  |  |  |  |
| Moderate ($20 \leq \text{score} \leq 46$) | | 0 (0) | 1 (14) | 1 (10) |
| Mild ($\text{score} \geq 47$) | | 3 (100) | 6 (86) | 9 (90) |
| FMA-LE |  | 28 [25–28] | 30 [28–31] | 28 [28–30] |
| ARAT |  | 31 [23–34] | 43 [28–50] | 34 [27–45] |
| BBT | Paralyzed side | 22 [12–25] | 25 [10–38] | 24 [7–28] |
|  | Non-paralyzed side | 53 [53–56] | 59 [53–61] | 57 [53–60] |
| mAs | 0 | 0 (0) | 0 (0) | 0 (0) |
|  | 1 | 2 (67) | 6 (86) | 8 (80) |
|  | 1+ | 1 (33) | 1 (14) | 2 (20) |
| BBS |  | 55 [54–55] | 56 [55–56] | 55 [55–56] |
| FRT (cm) |  | 29 [28–30] | 38 [33–40] | 33 [29–39] |
| SWT | Normal | 3 (100) | 4 (57) | 7 (70) |
|  | Decline | 0 (0) | 3 (43) | 3 (30) |
| TST | Normal | 3 (100) | 6 (86) | 9 (90) |
|  | Decline | 0 (0) | 1 (14) | 1 (10) |
| BI |  | 100 [100] | 100 [100] | 100 [100] |

Values are expressed as numbers (%) or medians [25<sup>th</sup>–75<sup>th</sup> percentile].
ARAT, action research arm test; BBS, Berg balance scale; BBT, Box and Block Test; BI, Barthel Index; BMI, body mass index; CI, cerebral infarction; FMA-LE, Fugl-Meyer assessment of the lower extremity; FMA-UE, Fugl-Meyer assessment of the upper extremity; FRT, functional reach test; ICH, intracranial hemorrhage; mAs, modified Ashworth scale; SWT, Semmes-Weinstein monofilament test; TST, thumb search test.

### Outcome data

Tables 2 and 3 present the results for the kinematic data on outward and inward reaching motions. Fig 5 shows the changes in the joint angles of shoulder flexion, shoulder abduction, and elbow flexion during the outward and return motions of the reaching motion.

**Fig 5.**
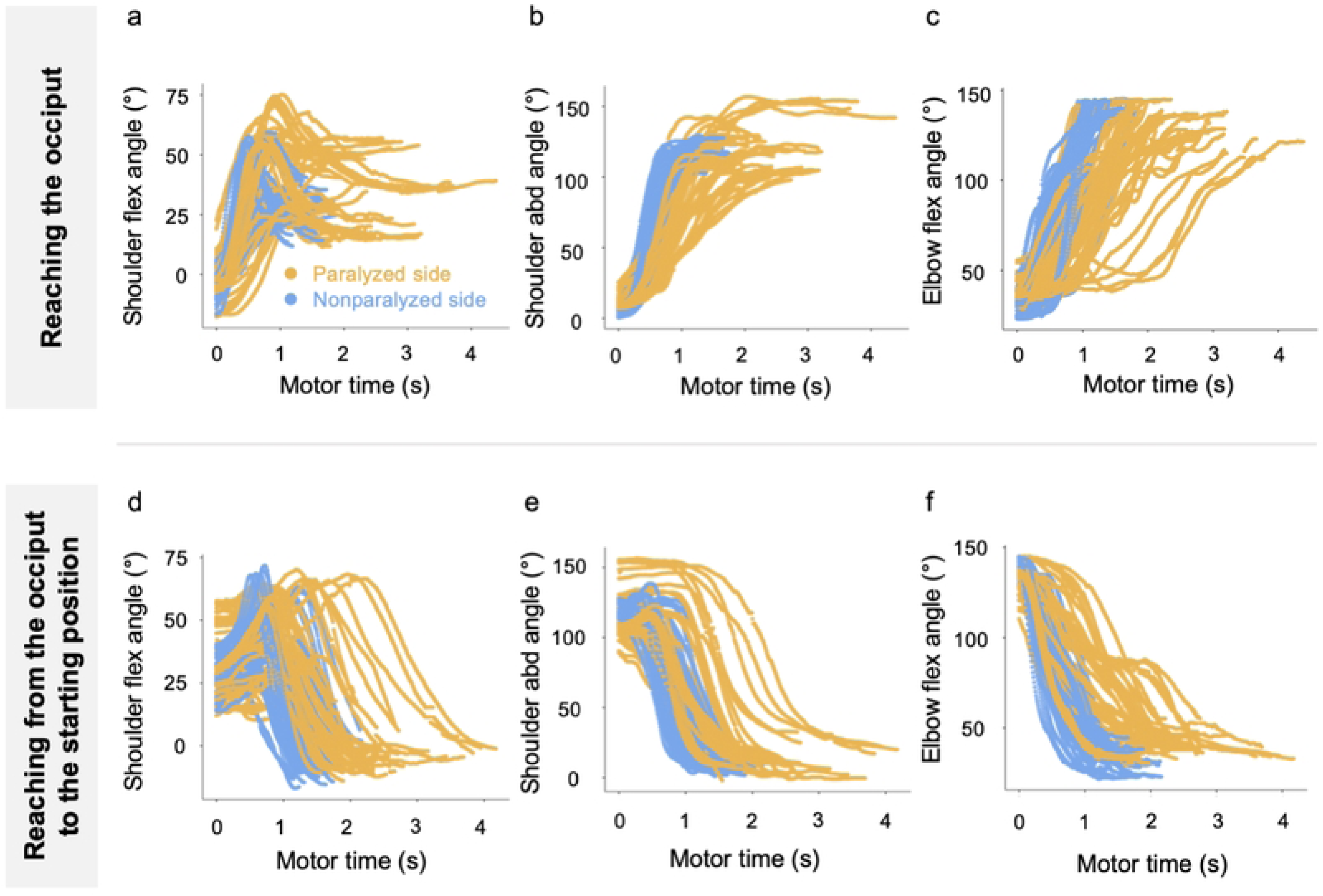
Changes in the joint angle during the reaching motion. (a) Changes in the shoulder flex angle, (b) changes in the shoulder abd angle, (c) changes in the elbow flex angle while reaching the occiput, (d) changes in the shoulder flex angle, (e) changes in the shoulder abd angle, and (f) changes in the elbow flex angle while reaching from the occiput to the starting limb position. abd, abduction; flex, flexion.

**Table 2.**
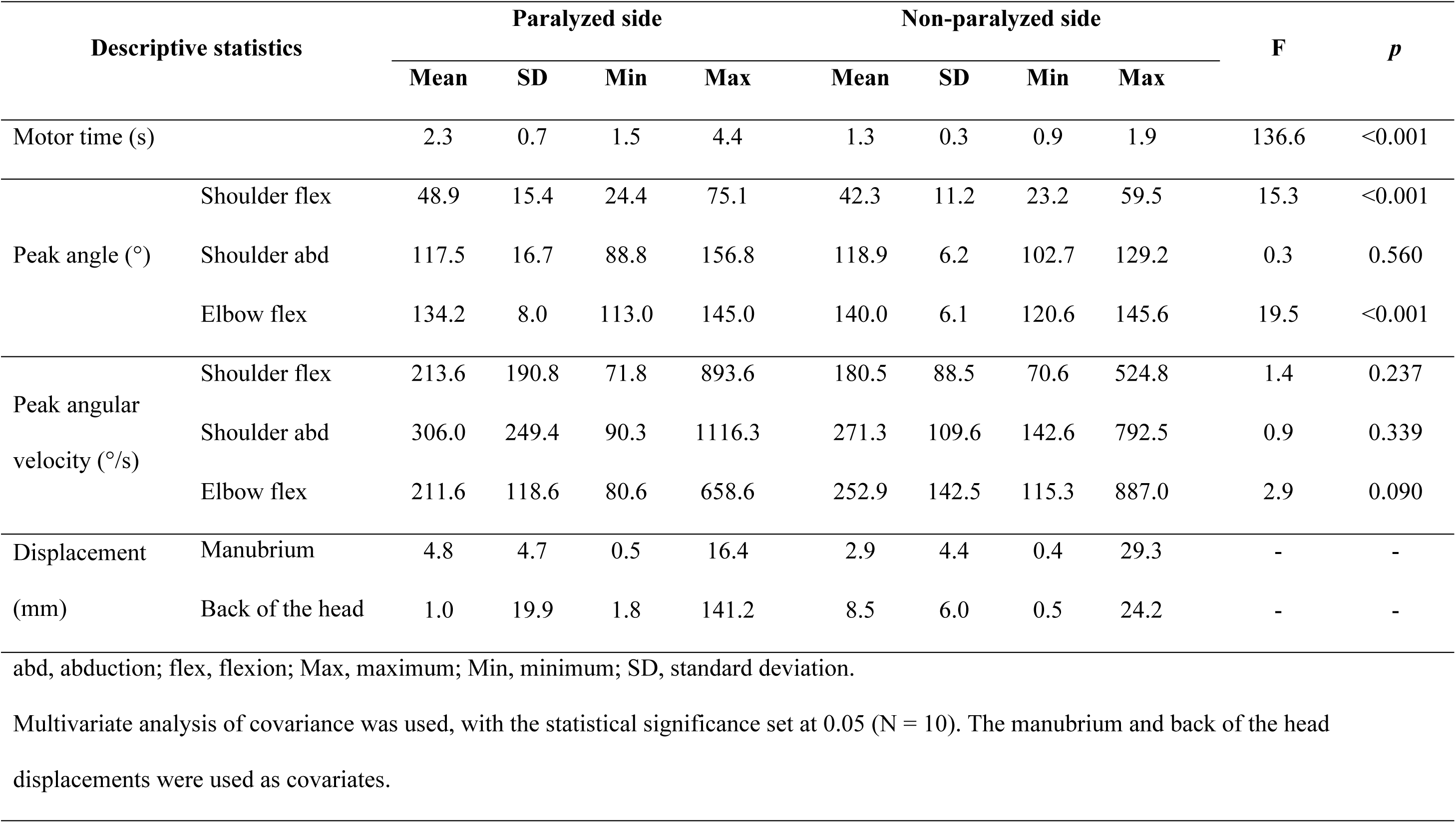
Outward motion of the reaching task: results of multivariate analysis of covariance.

**Table 3.**
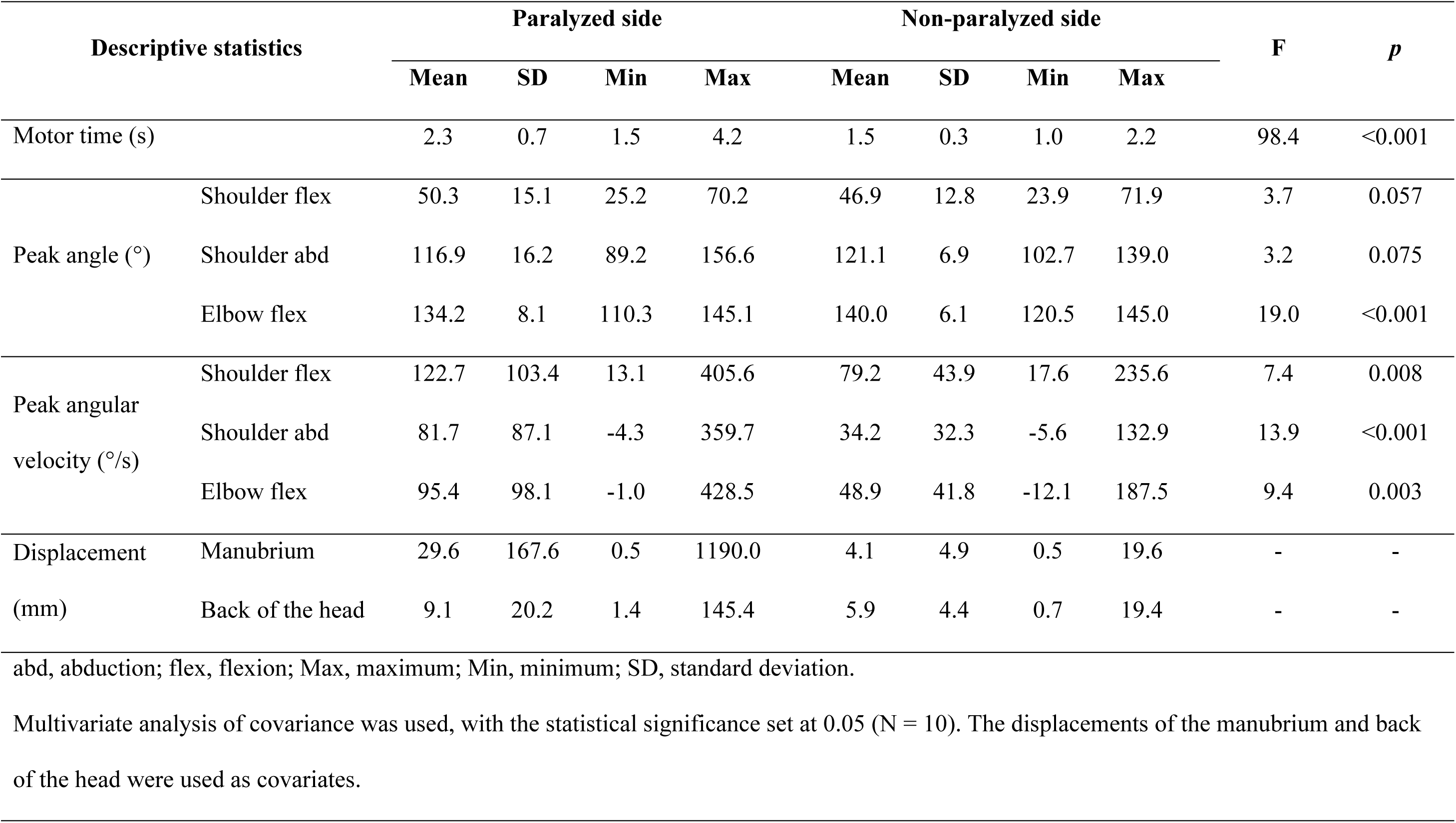
Return motion of the reaching task: results of multivariate analysis of covariance.

### Main findings

#### Detection of motion features

Multivariate analysis of covariance was performed. For the outward motion of the reaching task, Wilks’ lambda test showed a significant main effect (F [7, 86] = 25.1, *p* < 0.001) on the measurement side. The paralyzed side had a significantly longer motor time (F = 136.6, *p* < 0.001). The peak shoulder flexion angle was significantly greater on the paralyzed side (F = 15.3, *p* < 0.001), and the peak elbow flexion angle was significantly greater on the non-paralyzed side (F = 19.5, *p* < 0.001). There were no differences in the peak shoulder abduction angle (F = 0.3, *p* = 0.560) and peak angular velocities of shoulder flexion (F = 1.4, *p* = 0.237), shoulder abduction (F = 0.9, *p* = 0.339), and elbow flexion (F = 2.9, *p* = 0.090; Table 2). For the return motion of the reaching task, Wilks’ lambda test showed a significant main effect (F [7, 86] = 31.5, *p* < 0.001) on the measured side. Motor time (F = 98.4, *p* < 0.001) was significantly longer on the paralyzed side, as were the peak elbow flexion angle (F = 19.0, *p* < 0.001) and the peak angular rates of shoulder flexion (F = 7.4, *p* = 0.008), shoulder abduction (F = 13.9, *p* < 0.001), and elbow flexion (F = 9.4, *p* = 0.003). There was no significant difference in the peak shoulder flexion angle (F = 3.7, *p* = 0.057) or peak shoulder abduction angle (F = 3.2, *p* = 0.075; Table 3).

Binomial logistic regression analysis of the model-fitted features on the paralyzed and non-paralyzed sides was performed using multivariate analysis of covariance. During the outward motion, the regression model was fitted for the motor time (X^2^ = 112, *p* < 0.001), peak shoulder flexion angle (X^2^ = 18.4, *p* = 0.010), and peak elbow flexion angle (X^2^ = 24.9, *p* < 0.001; Table 4 and Fig 6). During the return motion, the regression model was fitted for the motor time (X^2^ = 87.5, *p* < 0.001), peak elbow flexion angle (X^2^ = 28.8, *p* < 0.001), peak angular velocity of shoulder abduction (X^2^ = 18.4, *p* = 0.01), and peak angular velocity of elbow flexion (X^2^ = 15.8, *p* = 0.03). The peak angular velocity of shoulder flexion (X^2^ = 11.8, *p* = 0.1) did not fit the regression model (Table 4 and Fig 7)

**Fig 6.**
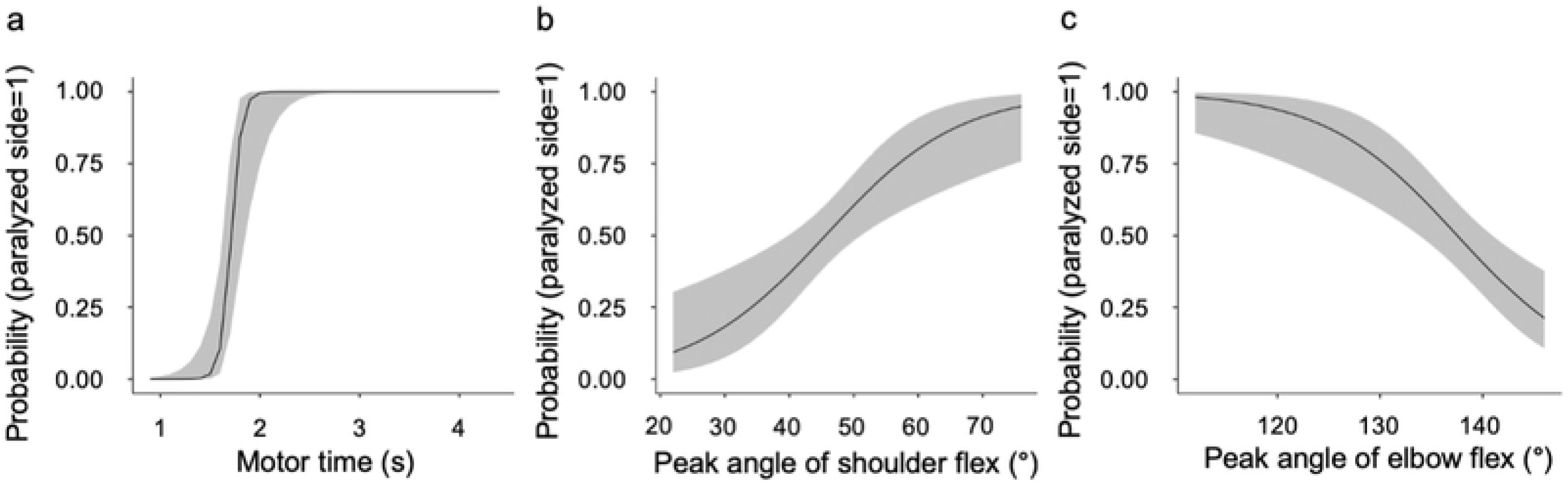
Results of the binomial logistic regression analysis for the features of reaching motion to the back of the head. (a) Motor time, (b) peak angle of shoulder flex, and (c) peak angle of elbow flex. flex, flexion.

**Fig 7.**
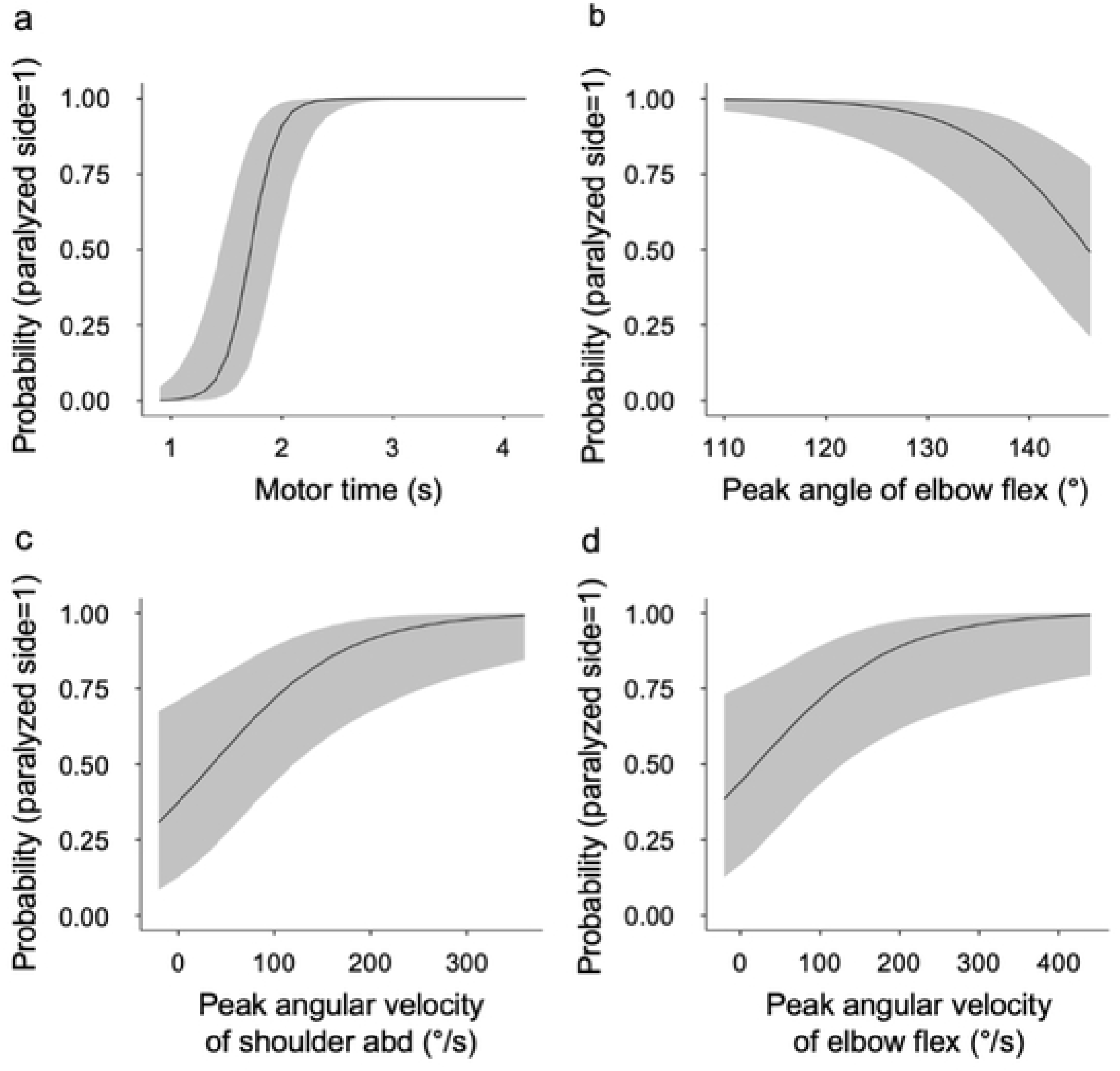
Results of the binomial logistic regression analysis for the features of reaching motion from the occiput to the starting position. (a) Motor time, (b) peak angle of elbow flex, (c) peak angular velocity of shoulder abd, and (d) peak angular velocity of elbow flex. abd, abduction; flex, flexion.

**Table 4.**
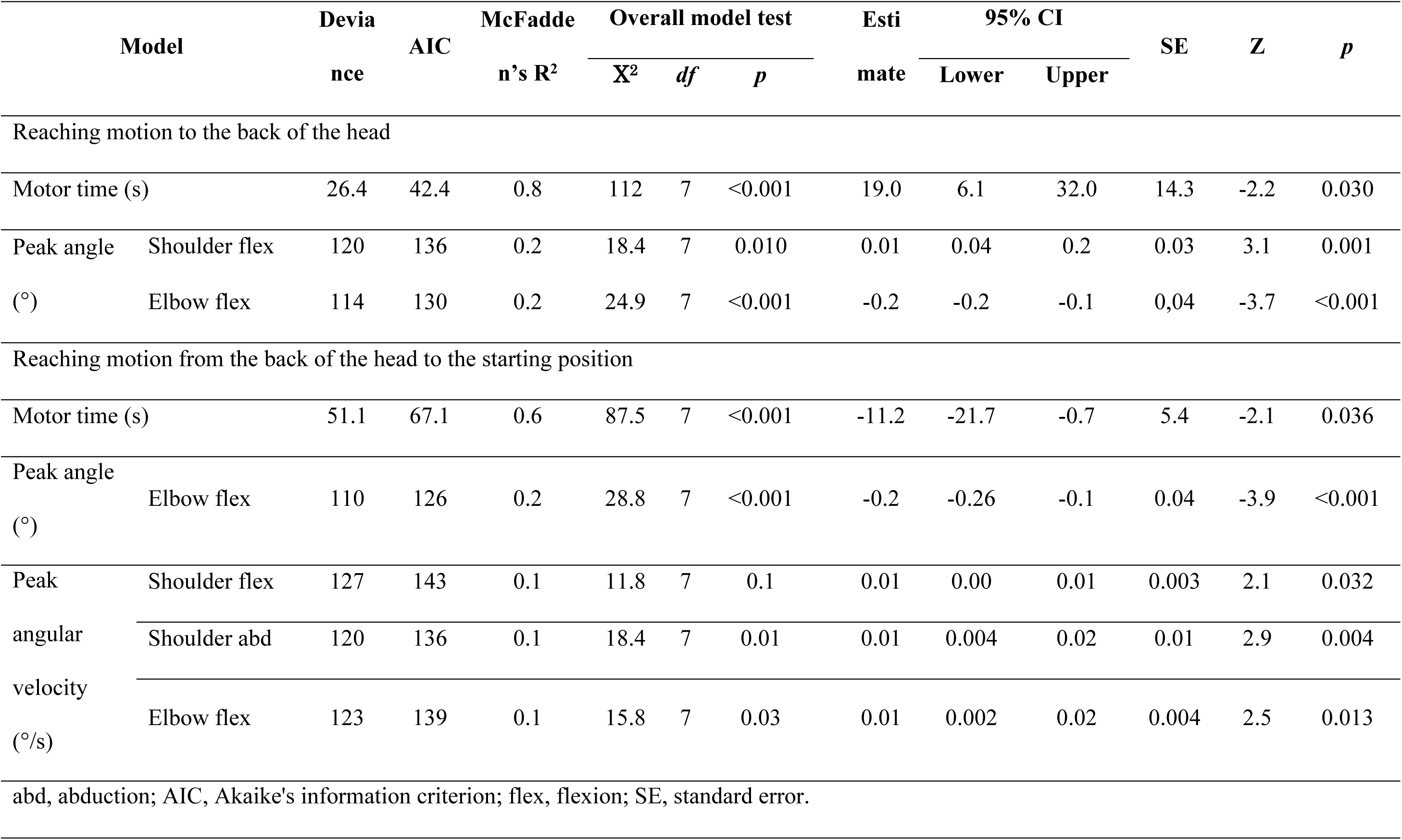
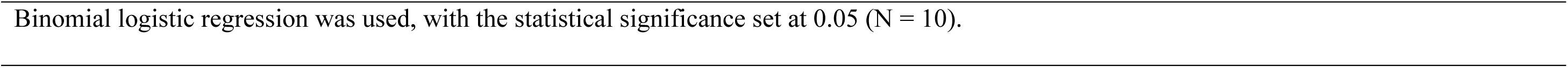
Model goodness of fit results of the binomial logistic regression analysis.

| Model |  | Deviance | AIC | McFadden's R<sup>2</sup> | Overall model test |  |  | Estimate | 95% CI |  | SE | Z | p |
| --- | --- | --- | --- | --- | --- | --- | --- | --- | --- | --- | --- | --- | --- |
|  |  |  |  |  | X<sup>2</sup> | df | p |  | Lower | Upper |  |  |  |
| Reaching motion to the back of the head |  |  |  |  |  |  |  |  |  |  |  |  |  |
| Motor time (s) |  | 26.4 | 42.4 | 0.8 | 112 | 7 | <0.001 | 19.0 | 6.1 | 32.0 | 14.3 | -2.2 | 0.030 |
| Peak angle | Shoulder flex | 120 | 136 | 0.2 | 18.4 | 7 | 0.010 | 0.01 | 0.04 | 0.2 | 0.03 | 3.1 | 0.001 |
| (°) | Elbow flex | 114 | 130 | 0.2 | 24.9 | 7 | <0.001 | -0.2 | -0.2 | -0.1 | 0.04 | -3.7 | <0.001 |
| Reaching motion from the back of the head to the starting position |  |  |  |  |  |  |  |  |  |  |  |  |  |
| Motor time (s) |  | 51.1 | 67.1 | 0.6 | 87.5 | 7 | <0.001 | -11.2 | -21.7 | -0.7 | 5.4 | -2.1 | 0.036 |
| Peak angle | Elbow flex | 110 | 126 | 0.2 | 28.8 | 7 | <0.001 | -0.2 | -0.26 | -0.1 | 0.04 | -3.9 | <0.001 |
| (°) |  |  |  |  |  |  |  |  |  |  |  |  |  |
| Peak | Shoulder flex | 127 | 143 | 0.1 | 11.8 | 7 | 0.1 | 0.01 | 0.00 | 0.01 | 0.003 | 2.1 | 0.032 |
| angular | Shoulder abd | 120 | 136 | 0.1 | 18.4 | 7 | 0.01 | 0.01 | 0.004 | 0.02 | 0.01 | 2.9 | 0.004 |
| velocity | Elbow flex | 123 | 139 | 0.1 | 15.8 | 7 | 0.03 | 0.01 | 0.002 | 0.02 | 0.004 | 2.5 | 0.013 |
| (°/s) |  |  |  |  |  |  |  |  |  |  |  |  |  |
| abd, abduction; AIC, Akaike's information criterion; flex, flexion; SE, standard error. |  |  |  |  |  |  |  |  |  |  |  |  |  |

Binomial logistic regression was used, with the statistical significance set at 0.05 (N = 10).

For features fitted to the regression model, cutoff values were calculated to discriminate between the paralyzed and non-paralyzed sides using the ROC curve. The cutoff values for the outward motion were 1.6 s for the motor time, 55° for the peak shoulder flexion angle, and 145° for the peak elbow flexion angle (Table 5). The cutoff values for the return motion were 1.6 s for the motor time, 145° for the maximum elbow flexion angle, 53°/s for the peak angular velocity of shoulder abduction, and 62°/s for the peak angular velocity of elbow flexion (Table 5). Among the cutoff values detected, the kinematic feature with the highest Youden’s index was the motor time for both outward and return motions, whereas the kinematic feature with the largest AUC was the motor time for both the outward motion (AUC = 0.96, SD = 0.02, *p* = 0.00) and return motion (AUC = 0.92, SD = 0.03, *p* = 0.00).

**Table 5.** Results of the receiver operating characteristic curve.

| Scale |  | Cutoff point | Sensitivity (%) | Specificity (%) | Youden's index | AUC |
| --- | --- | --- | --- | --- | --- | --- |
| Reaching the occiput |  |  |  |  |  |  |
| Motor time (s) |  | 1.6 | 80 | 98 | 0.78 | 0.96 |
| Peak angle (°) | Shoulder flex | 55 | 84 | 44 | 0.28 | 0.63 |
|  | Elbow flex | 145 | 96 | 4 | 0.00 | 0.28 |
| Reaching from the occiput to the starting limb position |  |  |  |  |  |  |
| Motor time (s) |  | 1.6 | 78 | 90 | 0.68 | 0.92 |
| Peak angle (°) | Elbow flex | 145 | 100 | 6 | 0.06 | 0.29 |
| Peak angular velocity (°/s) | Shoulder abd | 53 | 84 | 48 | 0.32 | 0.65 |
|  | Elbow flex | 62 | 74 | 56 | 0.30 | 0.67 |
| abd, abduction; AUC, area under the curve; flex, flexion. |  |  |  |  |  |  |
| Receiver operating characteristic curve was used. |  |  |  |  |  |  |

#### Pattern analysis of upper limb movements

The patterns of changes in the motor time and joint angle on the paralyzed and non-paralyzed sides of the reaching motion to the occipital region were analyzed using random forest clustering. Using the elbow method, the number of clusters was determined from the plotted values of the sum of the squares of the intra-cluster errors for each cluster (Fig 8). The number of clusters was determined to be four for the paralyzed side and three for the non-paralyzed side of the outward motion as well as four for the paralyzed and non-paralyzed sides of the return motion. The clustering structure for the four conditions is presented in Figs 9 and 10. As an example, in the results of the motor time on the paralyzed side of the reaching motion to the occiput, four clusters are illustrated. As the abscissa is the z-value, the motor time of cluster 1 was shorter than the cutoff value. By contrast, cluster 2 had motion times within and longer than the cutoff values, and clusters 3 and 4 had longer motion times than the cutoff values. For the outward motion, the number of clusters on the paralyzed side was four (N = 11,542; R^2^ = 0.40; AIC = 27,756; Bayesian information criterion [BIC] = 27,873) and that on the non-paralyzed side was three (N = 6648; R^2^ = 0.41; AIC = 15,673; BIC = 15,754). For the return motion, the number of clusters on the paralyzed side was four (N = 11,574; R^2^ = 0.34; AIC = 30,581; BIC = 30,699) and that on the non-paralyzed side was four (N = 7429; R^2^ = 0.50; AIC = 14974; BIC = 15,085; Table 6).

**Fig 8.**
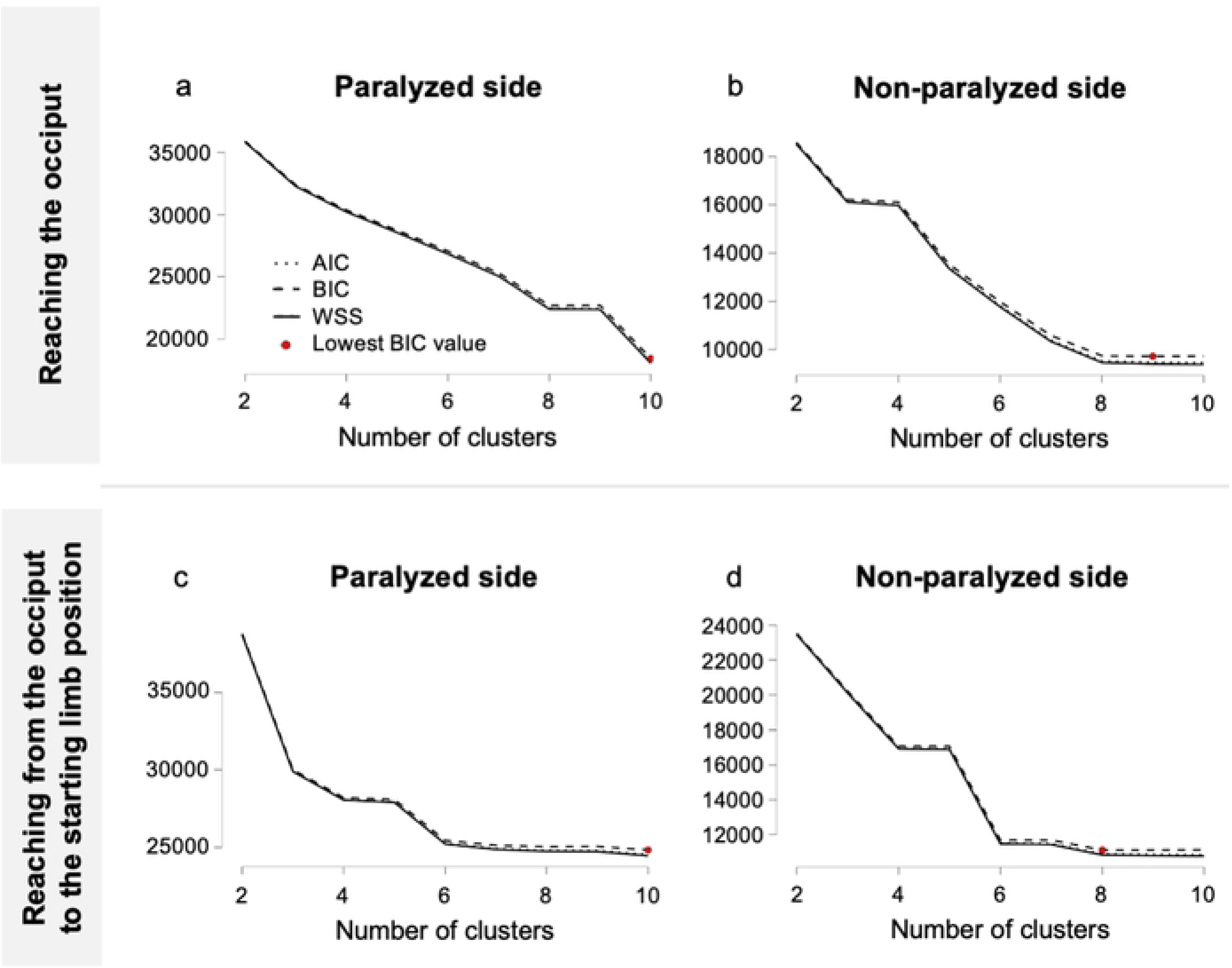
Random forest clustering of the elbow method plot. A plot of the intra-cluster sums of squares of errors for each cluster is shown in the figure. The number of clusters is (a) four for the paralyzed side and (b) three for the non-paralyzed side of the reaching motion to the occiput; and (c) four for the paralyzed side and (d) four for the non-paralyzed side of the reaching motion from the occiput to the starting position. AIC, Akaike’s information criterion; BIC, Bayesian information criterion; WSS, within sum of squares

**Fig 9.**
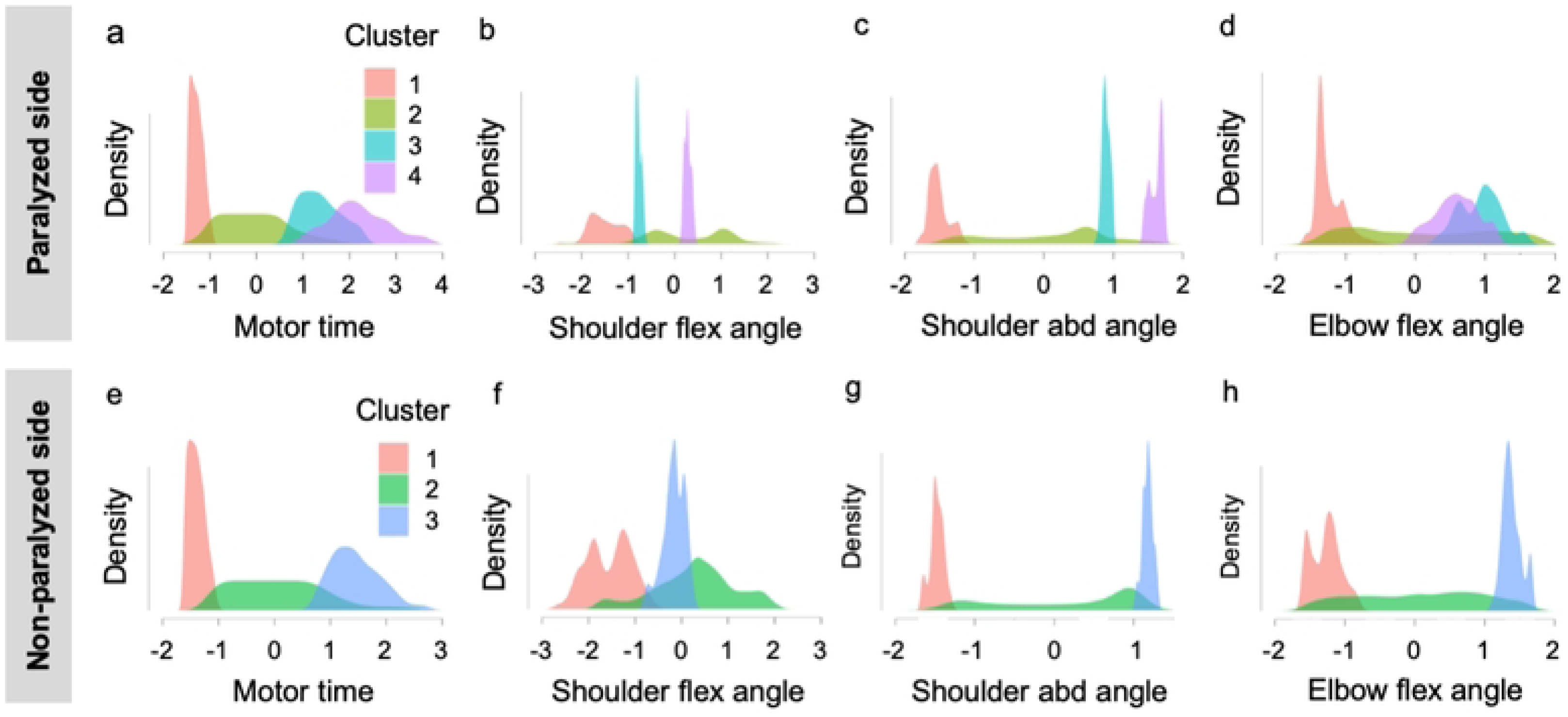
Random forest clustering of the features of reaching motion to the occiput. The upper panel shows the clustering structure of the (a) motor time, (b) shoulder flex angle, (c) shoulder abd angle, and (d) elbow flex angle on the paralyzed side. In the lower panel, the clustering structure of the (e) motor time, (f) shoulder flex angle, (g) shoulder abd angle, and (h) elbow flex angle for the non-paralyzed side is shown. The horizontal axis of the figure indicates the Z-values. The areas of each cluster composed of densities and absolute Z-values of each parameter are all equal. abd, abduction; flex, flexion.

**Fig 10.**
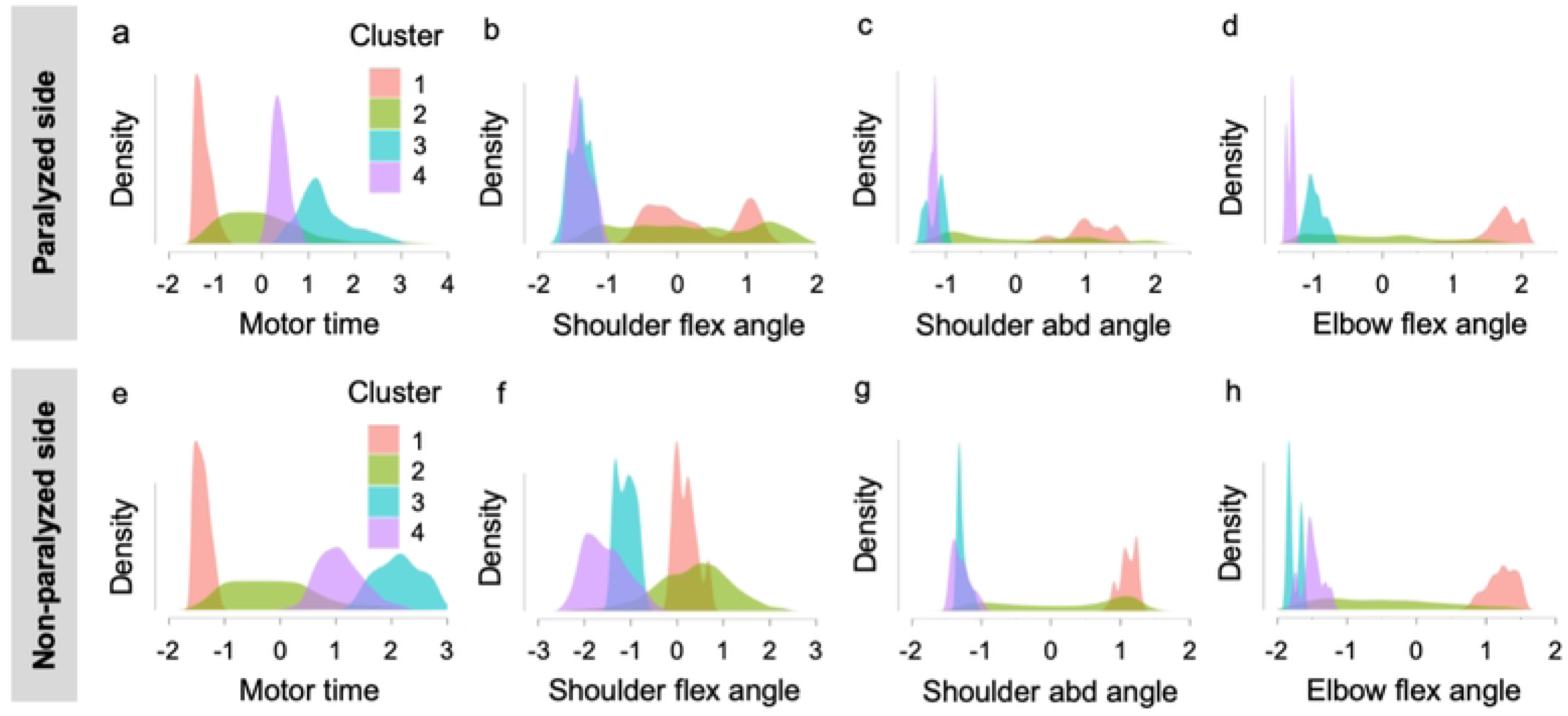
Random forest clustering of the features of reaching motion from the occiput to the starting position. The upper panel shows the clustering structure of the (a) motor time, (b) shoulder flex angle, (c) shoulder abd angle, and (d) elbow flex angle on the paralyzed side. In the lower panel, the clustering structure of the (a) motor time, (b) shoulder flex angle, (c) shoulder abd angle, and (d) elbow flex angle on the paralyzed side. In the lower panel, the clustering structure of the (e) motor time, (f) shoulder flex angle, (g) shoulder abd angle, and (h) elbow flex angle for the non-paralyzed side is shown. The horizontal axis of the figure indicates the Z-values. The areas of each cluster composed of densities and absolute Z-values of each parameter are all equal. abd, abduction; flex, flexion.

**Table 6.** Results of random forest clustering.

| Measuring side | Clusters | N | R<sup>2</sup> | AIC | BIC |
| --- | --- | --- | --- | --- | --- |
| Reaching motion to the occiput |  |  |  |  |  |
| Paralyzed side | 4 | 11542 | 0.40 | 27756.22 | 27873.88 |
| Non-paralyzed side | 3 | 6648 | 0.41 | 15673.33 | 15754.96 |
| Reaching motion from the occiput to the starting position |  |  |  |  |  |
| Paralyzed side | 4 | 11574 | 0.34 | 30581.45 | 30699.15 |
| Non-paralyzed side | 4 | 7429 | 0.50 | 14974.92 | 15085.53 |

AIC, Akaike's information criterion; BIC, Bayesian information criterion.

## Discussion

In this study, a multivariate analysis of covariance was conducted to test the hypothesis that the kinematic characteristics differ between the paralyzed and non-paralyzed upper limbs when reaching the occiput in patients with mild hemiplegia in the chronic phase. During the outward reaching motion, the paralyzed side showed a significantly longer motor time and significantly greater peak shoulder flexion angle. In contrast, the peak elbow flexion angle was significantly greater on the non-paralyzed side. During the return reaching motion, the motor time was significantly longer on the paralyzed side and the peak angular velocities of elbow flexion, shoulder flexion, shoulder abduction, and elbow flexion were significantly greater on the paralyzed side. Next, to detect the cutoff values of the motor features that discriminate between the paralyzed and non-paralyzed sides, binomial logistic regression analysis was conducted on the features fitted to the models for the paralyzed and non-paralyzed sides using multivariate analysis of covariance. For the features fitted to the regression model, the cutoff values discriminating between the paralyzed and non-paralyzed sides were calculated using the ROC curve. As a result, during the outward reaching motion, the motor time, peak angular velocity of shoulder flexion, and peak angular velocity of elbow flexion were calculated to be 1.6 s, 55°, and 145°, respectively. During the return reaching motion, the motor time, peak elbow joint flexion angle, peak angular velocity of shoulder abduction, and peak angular velocity of elbow flexion were calculated to be 1.6 s, 145°, 53°/s, and 62°/s, respectively. Therefore, the calculated cutoff values can be used as the target values for treatment to improve the reaching motion to the occiput in patients with mild hemiplegia.

Among the cutoff values detected, the kinematic feature with the highest Youden’s index was the motor time for both the outward and return reaching motions. Previous studies have reported that patients with stroke have a longer motor time, and the most difficult subitem in the FMA-UE is the motor time in Part D [47,48]. Motor time is presumed to be an index that determines whether the reaching motion to the occiput in a patient is a near-normal motion. In this study, the cutoff value for the peak elbow flexion angle was 145° for both the outward and return reaching motions. Hairdressing with a reaching motion to the back of the head requires the highest elbow joint flexion angle among the verified daily activities [49]. Hence, we inferred that sufficient elbow joint flexion is important to reach the occiput. In the outward reaching motion, the peak angle was significantly larger on the paralyzed side during shoulder flexion and on the non-paralyzed side during elbow flexion. Reaching the paralyzed side in patients with stroke has been reported to result in decreased coordination of the shoulder and elbow joint movements [50]. In the outward motion of reaching training, the goal of the exercise is to shorten the motor time and increase the elbow flexion angle more than the shoulder flexion angle. The cutoff values for angular acceleration of the joints were detected in the return reaching motion. When the hand reached the occiput, the upper limb was in abduction at the shoulder joint and flexion at the elbow joint, which is the joint motion pattern of the flexor muscles of the upper limb that appears after a stroke [51,52]. A previous study reported that muscle spasticity affects the angular acceleration of joints in reaching exercises using the paralyzed upper limb in patients with stroke [53]. The limb position, in which the hand reaches the occiput, induces a joint movement pattern of the upper limb and tends to increase the muscle tone of the flexor muscle group. Therefore, it may be difficult to control the angular velocity of the joint when the upper limb is lowered. In the return motion of reaching training, the practice goal is to shorten the movement time and decrease the angular velocity of shoulder abduction and elbow flexion.

In the present study, changes in the motion time and the joint angle of the paralyzed and non-paralyzed sides during the reaching motion to the occipital region were analyzed using random forest clustering. Using these results as a reference, one practical strategy is to approximate the cluster on the paralyzed side to the cluster on the non-paralyzed side of the patient. The motor time of cluster 1 on the paralyzed side in the reaching motion to the occiput was shorter than the cutoff value. The joint angles of cluster 1 in the paralyzed group were similar to those of cluster 1 in the non-paralyzed group. The motor time of cluster 2 on the paralyzed side was within and longer than the cutoff value. Cluster 2 on the paralyzed side exhibited a variety of joint angles similar to those of cluster 2 on the non-paralyzed side. For patients in cluster 2 on the paralyzed side whose motor time was approximately 1.6 s, we suggested that one of the exercises should be to shorten the motor time by reducing the variation in joint angles such that the hand can reach the occiput in the shortest distance. The motor times of patients in paralytic clusters 3 and 4 were longer than the cutoff values. Hence, we recommended that patients in paralytic clusters 3 and 4 practice joint movement patterns to approach the values of those in non-paralytic clusters 1 or 3. When targeting cluster 1 on the non-paralytic side, a method to reduce the changes in the joint angle and shorten the motor time exists. When targeting cluster 3 on the non-paralyzed side, patients in cluster 3 on the paralyzed side should be trained by increasing the angular changes in shoulder flexion and elbow flexion while aiming for a motor time near the cutoff value. For patients in cluster 4 on the paralyzed side, the motor time was aimed to be near the cutoff value, and the goals of practice were to decrease the angular change in shoulder abduction and increase the angular change in elbow flexion (S1 Table).

In the reaching motion from the occiput to the starting limb, the motion time of cluster 1 on the paralyzed side was shorter than the cutoff value, and, as in the reaching motion to the occiput, the motion pattern of cluster 1 on the paralyzed side was similar to that on the non-paralyzed side. The motor time of cluster 2 on the paralyzed side was within or near the cutoff value. For cluster 2 on the paralyzed side, one of the practice goals was to shorten the motor time. The motor times of clusters 3 and 4 on the paralyzed side were longer than the cutoff values. The joint angles of clusters 3 and 4 on the paralyzed side were similar to those of clusters 3 and 4 on the non-paralyzed side. These results suggest that there is no difference in the motion pattern between the paralyzed and non-paralyzed sides when reaching from the occiput to the starting position. For the reaching motion from the occiput to the starting position, we proposed a practice to control the angular velocity of shoulder abduction and elbow flexion, with the target motor time within or near the cutoff value (S1 Table).

This study had several limitations. First, the participants were patients with mild hemiplegia, whose condition persisted for more than 6 months; therefore, the obtained results should be used with caution in patients with early onset of the disease. Furthermore, eight of 10 participants in this study had a mAs score of 1 point, and we did not include patients with significant spasticity. Patients with severe motor paralysis are more likely to have strong muscle spasticity and exhibit joint movement patterns in the upper limbs [54]. Patients with severe motor paralysis who are unable to reach the occiput with their hands may have difficulties in practicing joint movement patterns based on the results of this study. Second, we did not compare the movements of the paralyzed upper limbs of patients with stroke with those of healthy participants. Patients with post-stroke hemiplegia also suffer from motor paralysis of the trunk and lower limb muscles, and their balance function may be impaired [55,56], and, during the reaching motion, the trunk and lower limb functions influence the upper limb joint motion [57,58]. Therefore, in this study, the sitting posture at the time of measurement was defined in detail and the displacement of the markers attached to the trunk was used as an adjustment variable in the statistical analysis. Based on the results of the FMA-LE, BBS, and FRT, the participants in this study had mild motor paralysis of the lower limbs and good trunk and balance function. However, how the lower limb and trunk movements as well as the balance strategies for sitting and holding positions were involved in upper limb movements requires further analysis. To target the upper limb movements of individuals with no motor paralysis, an analysis of the upper limb movements of healthy participants is required. Third, the relationship between the dominant hand and joint movement patterns was not examined. The right side was the dominant hand before the disease onset in all participants; however, the current dominant hand changed to the left in five participants. To verify the influence of the dominant hand, joint movement patterns should be compared under the same conditions as those of the dominant hand before the disease onset and at the time of measurement [59]. Fourth, the distal joints of the upper extremities were not analyzed. During the reaching motion, limitations in the joint range of motion in the forearm and wrist joints affect the shoulder joint motion [60]. In this study, the joint motions of the shoulder and elbow joints were analyzed; however, coordination with the distal joints of the upper limb was not verified. Fifth, the motor tasks were measured at patients’ comfortable speeds. When the movement is measured at a specified speed, it may cause changes in the joint motion and compensatory movement strategies are needed [61,62]. It remains unclear whether the practice of movement patterns performed in accordance with the optimal movement time and angular velocity on the non-paralyzed side can restore motor paralysis. Further validation is needed to consider these limitations to clinically utilize the practice methods and target values to reproduce movements equivalent to those of the non-paralyzed side without the appearance of spasticity or compensatory movements when upper limb motor paralysis is mild.

## Conclusions

In this study, the kinematic features and cutoff values while reaching the back of the head in patients with mild hemiplegia in the chronic phase were detected, and patterns of changes in the joint angle were analyzed. Based on our findings, when patients with hemiplegia who can reach the back of the head practice with the goal of smoother upper limb motion, the motion patterns of the non-paralyzed upper limb can be referenced to set the target values of motion time, joint angle, and angular velocity. The motor time of the paralyzed upper limb was measured, and the corresponding clusters were referred to from the results of random forest clustering. The results of this study will be used to plan motor practice strategies that trace the pattern of joint angles of the upper limb without paralysis. Moreover, the results obtained in this study can be used as a reference to devise effective practice methods to improve the reaching motion to the back of the head.

## Data Availability

Data cannot be shared publicly because of the Ethics Committee of the Jikei University School of Medicine. Data are only available upon request. Data are available from the Ethics Committee of the Jikei University School of Medicine for researchers who meet the criteria for access to confidential data.

## Acknowledgments

We would like to thank the occupational therapists at the Department of Rehabilitation Medicine, Jikei University Hospital, for their cooperation in obtaining the data for this study. Moreover, we would like to thank Prof. Michito Namekawa and Prof. Satoshi Kido, Department of Rehabilitation, Graduate School of Health Science, Saitama Prefectural University, for their clinical advice for the successful completion of this study.

## Author contributions

D.S., T.H. and M.A. collated the literature and conceived the study. D.S., T.H., N.K., T.Y., and Y.N. developed the protocol. D.S. recruited participants and conducted data collections. D.S., T.H., K.K., and R.S. were involved in data analysis. D.S. wrote the first draft of the manuscript. T.H., N.K., T.Y., Y.N., and M.A. reviewed and edited the manuscript. All authors have agreed with the submitted version of the manuscript.

## Supporting information

**S1 Table. Training methods devised from the pattern analysis results.**

## References

1. World Stroke Organization. Annual Report 2020. 2020 [accessed 2021 December 1]. Available from: https://www.world-stroke.org/about-wso/annual-reports

2. Langhorne P, Bernhardt J, Kwakkel G. Stroke rehabilitation. Lancet. 2011;377(9778):1693–1702. doi: 10.1016/S0140-6736(11)60325-5.

3. van Mierlo ML, van Heugten CM, Post MW, Hajós TR, Kappelle LJ, Visser-Meily JM. Quality of life during the first two years post stroke: the Restore4Stroke Cohort Study. Cerebrovasc Dis. 2016;41(1-2):19–26. doi: 10.1159/000441197.

4. Fugl-Meyer AR, Jääskö L. Post-stroke hemiplegia and ADL-performance. Scand J Rehabil Med Suppl. 1980;7:140–152.

5. Karger DW, Bayha F. Engineered work measurement: the principles, techniques and data of methods-time measurement, modern time and motion study, and related applications engineering data. 2nd ed. New York: Industrial Press, 1966.

6. World Health Organization. International Classification of Functioning, Disability and Health (ICF). 2001 [accessed 2021 December 1]. Available from: https://www.who.int/standards/classifications/international-classification-of-functioning-disability-and-health#:~:text=ICF%20is%20the%20WHO%20framework,and%20measure%20health%20and%20disability

7. Veeger HE, Magermans DJ, Nagels J, Chadwick EK, van der Helm FC. A kinematical analysis of the shoulder after arthroplasty during a hair combing task. Clin Biomech (Bristol, Avon). 2006;21 Suppl 1:S39–44. doi: 10.1016/j.clinbiomech.2005.09.012.

8. Magermans DJ, Chadwick EK, Veeger HE, van der Helm FC. Requirements for upper extremity motions during activities of daily living. Clin Biomech (Bristol, Avon). 2005;20(6):591–599. doi: 10.1016/j.clinbiomech.2005.02.006.

9. Nakamura R, Moriyama S, Yamada Y, Seki K. Recovery of impaired motor function of the upper extremity after stroke. Tohoku J Exp Med. 1992;168(1):11–20.doi: 10.1620/tjem.168.11.

10. Lyle RC. A performance test for assessment of upper limb function in physical rehabilitation treatment and research. Int J Rehabil Res. 1981;4(4):483–492. doi: 10.1097/00004356-198112000-00001.

11. De Souza LH, Hewer RL, Miller S. Assessment of recovery of arm control in hemiplegic stroke patients. 1. Arm function tests. Int Rehabil Med. 1980;2(1):3–9. doi: 10.3109/09638288009163947.

12. Bernshteĭn NA. The co-ordination and regulation of movements. 1st English ed. Oxford: Pergamon Press; 1967. p. 196.

13. Brunnstrom S. Motor testing procedures in hemiplegia: based on sequential recovery stages. Phys Ther. 1966;46(4):357–375. doi: 10.1093/ptj/46.4.357.

14. Alt Murphy M, Häger CK. Kinematic analysis of the upper extremity after stroke - how far have we reached and what have we grasped? Phys Ther Rev. 2015;20(3):137–155. doi: 10.1179/1743288X15Y.0000000002

15. Alt Murphy M, Willén C, Sunnerhagen KS. Responsiveness of upper extremity kinematic measures and clinical improvement during the first three months after stroke. Neurorehabil Neural Repair. 2013;27(9):844–853. doi: 10.1177/1545968313491008.

16. Patterson TS, Bishop MD, McGuirk TE, Sethi A, Richards LG. Reliability of upper extremity kinematics while performing different tasks in individuals with stroke. J Mot Behav. 2011;43(2):121–130. doi: 10.1080/00222895.2010.548422.

17. Wagner JM, Rhodes JA, Patten C. Reproducibility and minimal detectable change of three-dimensional kinematic analysis of reaching tasks in people with hemiparesis after stroke. Phys Ther. 2008;88(5):652–663. doi: 10.2522/ptj.20070255.

18. Caimmi M, Carda S, Giovanzana C, Maini ES, Sabatini AM, Smania N, et al. Using kinematic analysis to evaluate constraint-induced movement therapy in chronic stroke patients. Neurorehabil Neural Repair. 2008;22(1):31–39. doi: 10.1177/1545968307302923.

19. Schwarz A, Kanzler CM, Lambercy O, Luft AR, Veerbeek JM. Systematic review on kinematic assessments of upper limb movements after stroke. Stroke. 2019;50(3):718–727. doi: 10.1161/STROKEAHA.118.023531.

20. Chen YW, Liao WW, Chen CL, Wu CY. Kinematic descriptions of upper limb function using simulated tasks in activities of daily living after stroke. Hum Mov Sci. 2021;79:102834. doi: 10.1016/j.humov.2021.102834.

21. Kim K, Song WK, Lee J, Lee HY, Park DS, Ko BW, et al. Kinematic analysis of upper extremity movement during drinking in hemiplegic subjects. Clin Biomech (Bristol, Avon). 2014;29(3):248–256. doi: 10.1016/j.clinbiomech.2013.12.013.

22. van Andel CJ, Wolterbeek N, Doorenbosch CA, Veeger DH, Harlaar J. Complete 3D kinematics of upper extremity functional tasks. Gait Posture. 2008;27(1):120–127. doi: 10.1016/j.gaitpost.2007.03.002.

23. Kwakkel G, van Wegen EEH, Burridge JH, Winstein CJ, van Dokkum LEH, Alt Murphy M, et al. Standardized measurement of quality of upper limb movement after stroke: consensus-based core recommendations from the Second Stroke Recovery and Rehabilitation Roundtable. Neurorehabil Neural Repair. 2019;33(11):951–958. doi: 10.1177/1545968319886477.

24. de Paiva Silva FP, Freitas SM, Silva PV, Banjai RM, Alouche SR. Ipsilesional arm motor sequence performance after right and left hemisphere damage. J Mot Behav. 2014;46(6):407–414. doi: 10.1080/00222895.2014.924473.

25. Schwarz A, Veerbeek JM, Held JPO, Buurke JH, Luft AR. Measures of interjoint coordination post-stroke across different upper limb movement tasks. Front Bioeng Biotechnol. 2020;8:620805. doi: 10.3389/fbioe.2020.620805.

26. Woodbury ML, Velozo CA, Richards LG, Duncan PW. Rasch analysis staging methodology to classify upper extremity movement impairment after stroke. Arch Phys Med Rehabil. 2013;94(8):1527–1533. doi: 10.1016/j.apmr.2013.03.007.

27. Cirstea MC, Mitnitski AB, Feldman AG, Levin MF. Interjoint coordination dynamics during reaching in stroke. Exp Brain Res. 2003;151(3):289–300. doi: 10.1007/s00221-003-1438-0.

28. Yuji K, Makoto S, Akihisa O, Kazuhisa T, Yasuhiro T, Toyohiro H. Distinction of students and expert therapists based on therapeutic motions on a robotic device using support vector machine. J Med Biol Eng. 2020;40:790–797. doi: 10.1007/s40846-020-00562-3

29. Limited VMS. Plug-in Gait reference guide. 2021 [accessed 2021 December 1]. Available from: https://docs.vicon.com/display/Nexus212/PDF+downloads+for+Vicon+Nexus

30. Fugl-Meyer AR, Jääskö L, Leyman I, Olsson S, Steglind S. The post-stroke hemiplegic patient. 1. a method for evaluation of physical performance. Scand J Rehabil Med. 1975;7(1):13–31. doi: 10.2340/1650197771331

31. Carroll D. A quantitative test of upper extremity function. J Chronic Dis. 1965;18:479–491. doi: 10.1016/0021-9681(65)90030-5.

32. Desrosiers J, Bravo G, Hébert R, Dutil E, Mercier L. Validation of the Box and Block Test as a measure of dexterity of elderly people: reliability, validity, and norms studies. Arch Phys Med Rehabil. 1994;75(7):751–755. doi: 10.1016/0003-9993(94)90130-9

33. Mathiowetz V, Volland G, Kashman N, Weber K. Adult norms for the Box and Block Test of manual dexterity. Am J Occup Ther. 1985;39(6):386–391. doi: 10.5014/ajot.39.6.386.

34. Bohannon RW, Smith MB. Interrater reliability of a modified Ashworth scale of muscle spasticity. Phys Ther. 1987;67(2):206–207. doi: 10.1093/ptj/67.2.206.

35. Berg KO, Wood-Dauphinee SL, Williams JI, Maki B. Measuring balance in the elderly: validation of an instrument. Can J Public Health. 1992;83 Suppl 2:S7–11.

36. Berg K. Measuring balance in the elderly: preliminary development of an instrument. Physiother Can. 1989;41(6):304–311. doi: 10.3138/ptc.41.6.304

37. Duncan PW, Weiner DK, Chandler J, Studenski S. Functional reach: a new clinical measure of balance. J Gerontol. 1990;45(6):M192–7. doi: 10.1093/geronj/45.6.m192.

38. Bell-Krotoski J, Tomancik E. The repeatability of testing with Semmes-Weinstein monofilaments. J Hand Surg Am. 1987;12(1):155–161. doi: 10.1016/s0363-5023(87)80189-2.

39. Hirai K. Maternal finger search test - examination of joint localization disorder (in Japanese). Clin Neurol. 1986;26:448–454.

40. Mahoney FI, Barthel DW. Functional evaluation: the Barthel Index. Md State Med J. 1965;14:61–65. doi: 10.1037/t02366-000

41. Oldfield RC. The assessment and analysis of handedness: the Edinburgh inventory. Neuropsychologia. 1971;9(1):97–113. doi: 10.1016/0028-3932(71)90067-4.

42. DeLong ER, DeLong DM, Clarke-Pearson DL. Comparing the areas under two or more correlated receiver operating characteristic curves: a nonparametric approach. Biometrics. 1988;44(3):837–845. doi: 10.2307/2531595

43. Tibshirani R, Walther G, Hastie T. Estimating the number of clusters in a data set via the gap statistic. J R Stat Soc Series B Stat Methodol. 2001;63(2):411–423. doi: 10.1111/1467-9868.00293

44. Breiman L. Random forests. Mach Learn. 2001;45:5–32. doi: 10.1023/A:1010933404324

45. Schwarz G. Estimating the dimension of a model. Ann Stat. 1978;6(2):461–464. doi: 10.1214/aos/1176344136

46. Thorndike RL. Who belongs in the family? Psychometrika. 1953;18:267–276. doi: 10.1007/BF02289263

47. Tauchi Y, Kyougoku M, Takahashi K, Okita Y, Takebayashi T. Dimensionality and item-difficulty hierarchy of the Fugl-Meyer assessment of the upper extremity among Japanese patients who have experienced stroke. Top Stroke Rehabil. 2022;29(8):579–587. doi: 10.1080/10749357.2021.1965797.

48. Collins KC, Kennedy NC, Clark A, Pomeroy VM. Getting a kinematic handle on reach- to-grasp: a meta-analysis. Physiotherapy. 2018;104(2):153–166. doi: 10.1016/j.physio.2017.10.002.

49. Morrey BF, Askew LJ, Chao EY. A biomechanical study of normal functional elbow motion. J Bone Joint Surg Am. 1981;63(6):872–877. doi: 10.2106/00004623-198163060-00002

50. Schwarz A, Bhagubai MMC, Nies SHG, Held JPO, Veltink PH, Buurke JH, et al. Characterization of stroke-related upper limb motor impairments across various upper limb activities by use of kinematic core set measures. J Neuroeng Rehabil. 2022;19(1):2. doi: 10.1186/s12984-021-00979-0.

51. Roh J, Rymer WZ, Beer RF. Evidence for altered upper extremity muscle synergies in chronic stroke survivors with mild and moderate impairment. Front Hum Neurosci. 2015;9:6. doi: 10.3389/fnhum.2015.00006.

52. Roh J, Rymer WZ, Perreault EJ, Yoo SB, Beer RF. Alterations in upper limb muscle synergy structure in chronic stroke survivors. J Neurophysiol. 2013;109(3):768–781. doi: 10.1152/jn.00670.2012.

53. Lackritz H, Parmet Y, Frenkel-Toledo S, Baniña MC, Soroker N, Solomon JM, et al. Effect of post-stroke spasticity on voluntary movement of the upper limb. J Neuroeng Rehabil. 2021;18(1):81. doi: 10.1186/s12984-021-00876-6.

54. Hijikata N, Kawakami M, Ishii R, Tsuzuki K, Nakamura T, Okuyama K, et al. Item difficulty of Fugl-Meyer assessment for upper extremity in persons with chronic stroke with moderate-to-severe upper limb impairment. Front Neurol. 2020;11:577855. doi: 10.3389/fneur.2020.577855.

55. Hugues A, Di Marco J, Ribault S, Ardaillon H, Janiaud P, Xue Y, et al. Limited evidence of physical therapy on balance after stroke: A systematic review and meta-analysis. PLoS One. 2019;14(8):e0221700. doi: 10.1371/journal.pone.0221700.

56. Sánchez N, Acosta AM, Lopez-Rosado R, Stienen AHA, Dewald JPA. Lower extremity motor impairments in ambulatory chronic hemiparetic stroke: evidence for lower extremity weakness and abnormal muscle and joint torque coupling patterns. Neurorehabil Neural Repair. 2017;31(9):814–826. doi: 10.1177/1545968317721974.

57. Tomita Y, Mullick AA, Levin MF. Reduced kinematic redundancy and motor equivalence during whole-body reaching in individuals with chronic stroke. Neurorehabil Neural Repair. 2018;32(2):175–186. doi: 10.1177/1545968318760725.

58. Tomita Y, Feldman AG, Levin MF. Referent control and motor equivalence of reaching from standing. J Neurophysiol. 2017;117(1):303–315. doi: 10.1152/jn.00292.2016.

59. Xiao X, Hu HJ, Li LF, Li L. Comparison of dominant hand to non-dominant hand in conduction of reaching task from 3D kinematic data: Trade-off between successful rate and movement efficiency. Math Biosci Eng. 2019;16(3):1611–1624. doi: 10.3934/mbe.2019077.

60. Gates DH, Walters LS, Cowley J, Wilken JM, Resnik L. Range of motion requirements for upper-limb activities of daily living. Am J Occup Ther. 2016;70(1):7001350010p1- 7001350010p10. doi: 10.5014/ajot.2016.015487.

61. Mandon L, Boudarham J, Robertson J, Bensmail D, Roche N, Roby-Brami A. Faster reaching in chronic spastic stroke patients comes at the expense of arm-trunk coordination. Neurorehabil Neural Repair. 2016;30(3):209–220. doi: 10.1177/1545968315591704.

62. DeJong SL, Schaefer SY, Lang CE. Need for speed: better movement quality during faster task performance after stroke. Neurorehabil Neural Repair. 2012;26(4):362–373. doi: 10.1177/1545968311425926.

